# Predicting work disability among people with chronic conditions: a large prospective cohort study

**DOI:** 10.1101/2022.08.18.22278925

**Authors:** Solja T. Nyberg, Jaakko Airaksinen, Jaana Pentti, Jenni Ervasti, Markus Jokela, Jussi Vahtera, Marianna Virtanen, Marko Elovainio, G. David Batty, Mika Kivimäki

**Author notes:** **Correspondence to:** Dr. Solja T Nyberg, University of Helsinki, Clinicum, Faculty of Medicine, Tukholmankatu 8B, FI-00014 Helsingin yliopisto, Finland.

## Abstract

Few risk prediction scores are available to identify people at increased risk of work disability, particularly for those with an existing morbidity. We examined the predictive performance of disability risk scores for employees with chronic disease. We used prospective data from 88,521 employed participants (mean age 43.1) in the Finnish Public Sector Study which included people with chronic disorders: musculoskeletal disorder, depression, migraine, respiratory disease, hypertension, cancer, coronary heart disease, diabetes and comorbid depression and cardiometabolic disease. 105 predictors were assessed at baseline and participants were linked to a national disability pension register. During a mean follow-up of 8.6 years, 6836 (7.7%) participants got a disability pension, the incidence varying between 9.9% among participants with migraine and 27.7% in those with comorbid depression and cardiometabolic disease. C-statistics for an 8-item risk score, comprising age, self-rated health, number of sickness absences, socioeconomic position, number of chronic illnesses, sleep problems, BMI, and smoking at baseline, was 0.80 (95%CI: 0.80-0.81) for musculoskeletal disorders (N=33,601), 0.83 (0.82-0.84) for migraine (N=22,065), 0.82 (0.81-0.83) for respiratory disease (N=15,372) and exceeded 0.72 for other disease groups. With 30% estimated risk as a threshold, a positive test detection rate and false positive rate ranged from 42.2% and 18.8% (cancer) to 79.8% and 45.2% (comorbid depression and cardiometabolic disease). Predictive performance was not improved in models with a new set of predictors or re-estimated coefficients. In conclusion, the 8-item work disability risk score may serve as a scalable screening tool in identifying individuals with increased risk for work disability.

## Introduction

Current estimates suggest that over a billion people globally live with disability and 80% of these are of working age.[1] Work disability reduces quality of life and career opportunities and is a major economic problem for societies with growing old-age dependence ratio.[2] The costs of lost work in people with disabilities has been estimated to be as high as 7% of gross domestic product[3] and is predicted to increase. There is therefore a need to better identify at risk individuals if premature labour market exit is to be prevented.

While several studies have investigated predictors of work disability in the general working population and in groups with specific diseases, or those that have undergone treatment procedures,[4-11] few have examined prediction in high-risk individuals, such as those with common chronic conditions that increase the likelihood of work disability. For example, there are no well-validated and easily administered prediction tools available to determine the risk among employees who have depression, musculoskeletal disorders, respiratory disease, hypertension or cardiometabolic multimorbidity. This is an important limitation which may hinder cost-effective allocation of preventative measures and targeting of interventions to those who would benefit most.

We have previously formulated a risk prediction model for work disability for use in the general working population.[4] We now examine whether the variables used also have the capacity to accurately determine work disability risk among employees with chronic conditions. We first evaluated the predictive performance of this existing 8-item risk score[4] in nine common disease groups; we then developed a modified version where we re-estimate coefficients; and, lastly, build a model with a new set of predictors selected from a large pool of additional variables and ascertain whether these new models improved risk prediction. To evaluate the relevance of the risk models in clinical decision making, we dichotomized the score to distinguish test positives from test negatives, and examined detection rate and false positive rate for this measure.[12]

## Methods

### Study population

Participants were from the Finnish Public Sector study, a prospective cohort study of public sector employees from 10 municipalities and 21 hospitals in the same geographical areas in Finland. Participants responded to questionnaire surveys conducted in 2000-2002, 2004, 2008, and 2012. We used data from the full study population including employees with and without chronic conditions. Using self-reports of physician-diagnosed diseases and records from the cancer registry, we also categorized participants into subgroups with different chronic conditions including those living with musculoskeletal disorder, depression, migraine, respiratory disease, hypertension, cancer, coronary heart disease, diabetes and comorbid depression and cardiometabolic disease (co-occurrence of mental and physical illnesses with major public health importance). The survey when a particular condition was first reported was considered as the baseline for each participant. Ethical approval was obtained from the ethics committee of the Helsinki-Uusimaa Hospital District Ethics Committee (HUS/1210/2016). Details of the study design and participants have been previously described.[13]

### Potential risk predictors

We used a pool of 105 variables including those in the existing 8-item risk prediction model of work disability for the general working population: age, BMI, socioeconomic status (SES), smoking, number of chronic diseases, self-rated health, difficulty falling asleep and number of sickness absences in previous year before baseline.[4] Other available variables included sex, alcohol consumption, physical inactivity, psychological distress and working conditions (job control, job demands, job strain, effort, reward and effort-reward imbalance, relational justice, procedural justice, participatory safety, support for innovation, vision, task orientation, social capital at work place, shift work and working night shifts). The description assessment and categorisation of the potential predictors is provided in appendix (p. 2-4).

### Ascertainment of work disability

All study members were insured in some pension scheme. Records were obtained from the national register at the Finnish Centre of Pensions,[14] an organisation which has a statutory obligation to curate records of all pensions in Finland. Disability pension records including start date and diagnosis according to the World Health Organization International Classification of Diseases, version 10. Our primary outcome was full-time disability pensions (temporary or permanent), and is defined as the capacity for work being impaired by at least 60% due to a disease, injury, or other disability. These records have been widely used in other research contexts.[15-17].

### Statistical methods

We imputed missing data on predictors (3.7% of all observations) as follows: we first set missing responses on chronic diseases as “no” answers, missing number of cigarettes as zero for non-smokers, and height as median value of all non-missing responses per individual. Other predictors were imputed using single imputation with predictive mean matching.[18]

The follow-up ended in case of death, work disability, or a maximum follow-up time of 10 years. To examine whether the original findings from the general working population were replicable in our dataset, we computed C-statistics for the entire study population with and without chronic conditions and among those with no history of sickness absence one year prior to baseline (the low-risk population). We then defined three steps to select the best model for each disease group. In the first, we examined whether the existing model was valid within the disease groups, estimating work disability risk as:

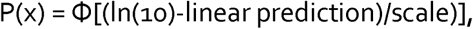

where Φ is the standard cumulative normal distribution. Coefficients for formulating linear predictor in the existing model are provided in appendix (page 5).

The second step was to examine whether the model could be improved by re-estimating the coefficients of the existing model. We used a similar method for fitting the models to that used in creating the original.[4] The third step was to examine whether the model could be improved by selecting a completely new set of predictors. For each disease group we first ran a redundancy analysis to exclude variables that could be readily predicted using all other variables. We then specified a parametric survival model that included all the candidate variables as predictors (‘full model’). To obtain a more parsimonious algorithm, we derived the predicted work disability risks from the full model for each individual. We then used backward stepwise ordinary least squares regression to select eight predictors, by predicting risks derived from the full model as the outcome. If the selected eight predictors included any summary variables (eg job strain), we repeated the previous steps with the summary variable(s) broken down to individual items.

The performance of each prediction model was evaluated using Harrell’s C-index, which is the concordance between predicted and observed survival.[18] Calibration of the model – that is, how accurately the predicted absolute risks correspond to the observed absolute risks – was assessed using calibration plots. We additionally plotted observed and predicted events by deciles of 10-year risk for each model, excluding by age and baseline those with less than 10 years of potential follow-up time. We compared the performance of the existing model with both new models using the C-statistics with 95% confidence intervals. If the confidence intervals were overlapping, the existing model was chosen.

To evaluate the final model for an individual, we dichotomized the score into ‘test positive’ versus ‘test negative’ using alternative cut-offs for risk scores. For positive test cases, we calculated false positive rate (the proportion of test positive cases who did not experience work disability), detection rate (the proportion of work disability cases who were test positive) and the ratio of true to false positives. The formulas were as follows:

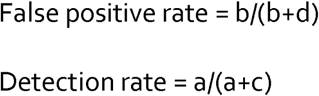

Ratio of true to false positives = 1 : (b/a), where a, b, c and d represent different combinations of risk scores and work disability as defined below:

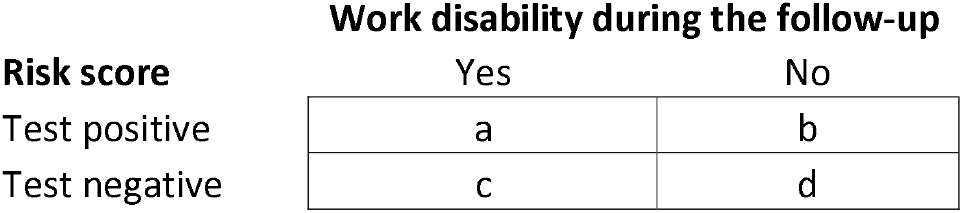

All analyses were performed using RStudio Version 1.4.1103 (packages: mice, rms and Hmisc) and SAS 9.4.

## Results

**Figure 1** shows sample selection. A total of 133 966 individuals (103 868 women) were mailed a self-administered questionnaire in 2000, 2004, 2008 or 2012. A total of 105 390 responded to at least one survey and, of these, 89 543 were linked to the national registry for work disability. We then excluded people who were on disability pension or retired, at age 65 years or older or with extreme values in BMI (<15 or >50) at baseline. In the analytical sample of 88 521 (70 805 women), the mean age of the participants was 43.1 years. Of the participants, 73 996 had no history of short-term work disability (no sickness absences one year before baseline) and this was denoted as the low-risk population. The number of participants with specific, prevalent condition at baseline varied between 1162 (comorbid depression and cardiometabolic disease) and 33 601 (musculoskeletal disorders). **Table 1** shows participant characteristics by chronic condition.

**Table 1.**
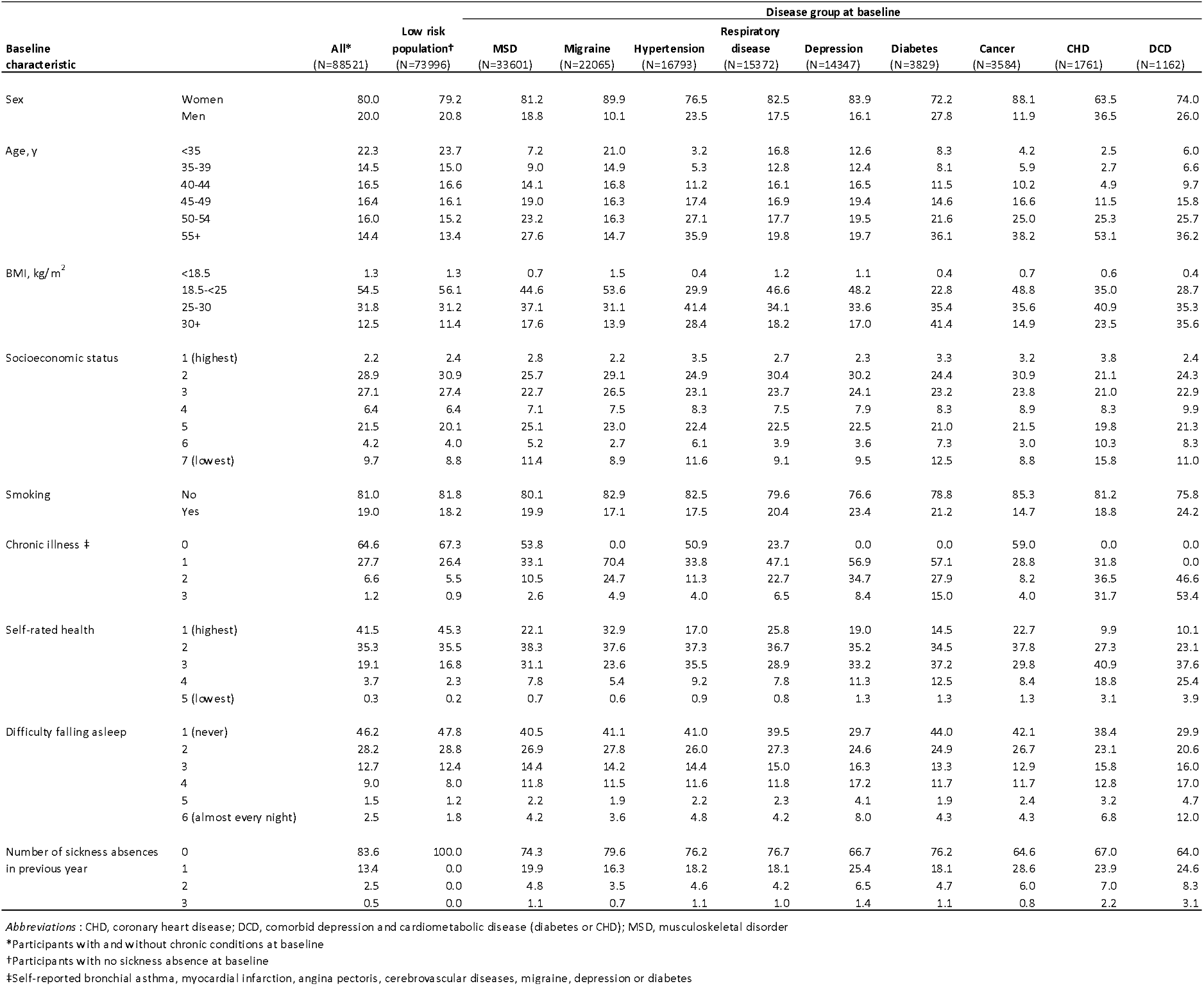
Baseline characteristics of all participants and subgroups of individuals with no history of sickness absence and those with a chronic condition at baseline

**Figure 1.**
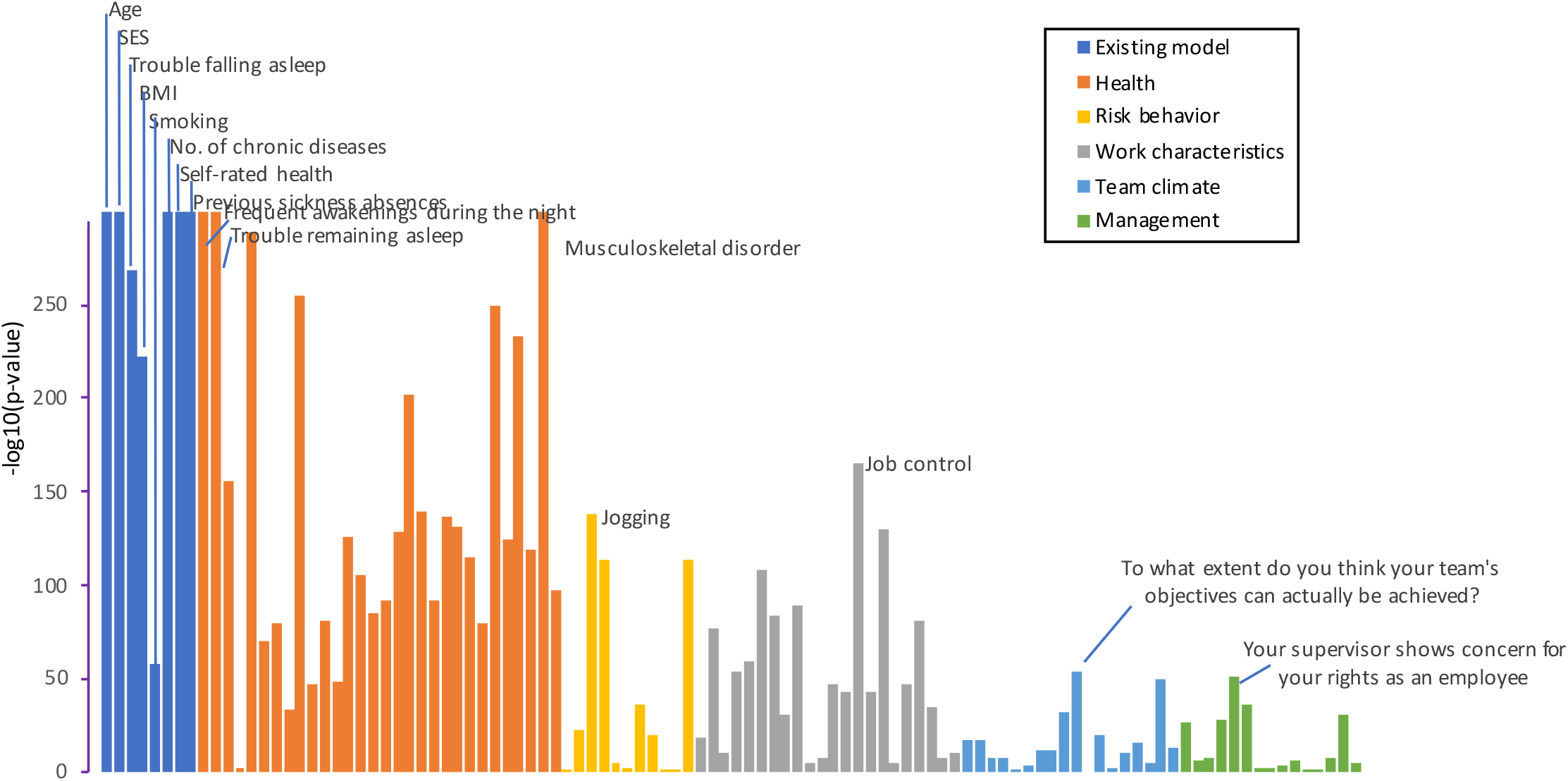
Study profile

During a mean follow-up of 8.6 years, 6836 (7.7%) participants were granted a disability pension. The incidence of work disability was 14.1% in those with musculoskeletal disorders, 9.9% for migraine, 15.1% for hypertension, 12.2% for respiratory disease (chronic bronchitis or asthma), 16.7% for depression, 16.4% for diabetes, 14.2% for cancer, 22.6% for coronary heart disease, and 27.7% for comorbid depression and cardiometabolic disease. The most common causes of work disability were diseases of the musculoskeletal system and connective tissue (44.2%), followed by mental, behavioural and neurodevelopmental disorders (23.7%), and these proportions varied by disease group (**figure 2**, for details, see appendix p. 6).

**Figure 2.**
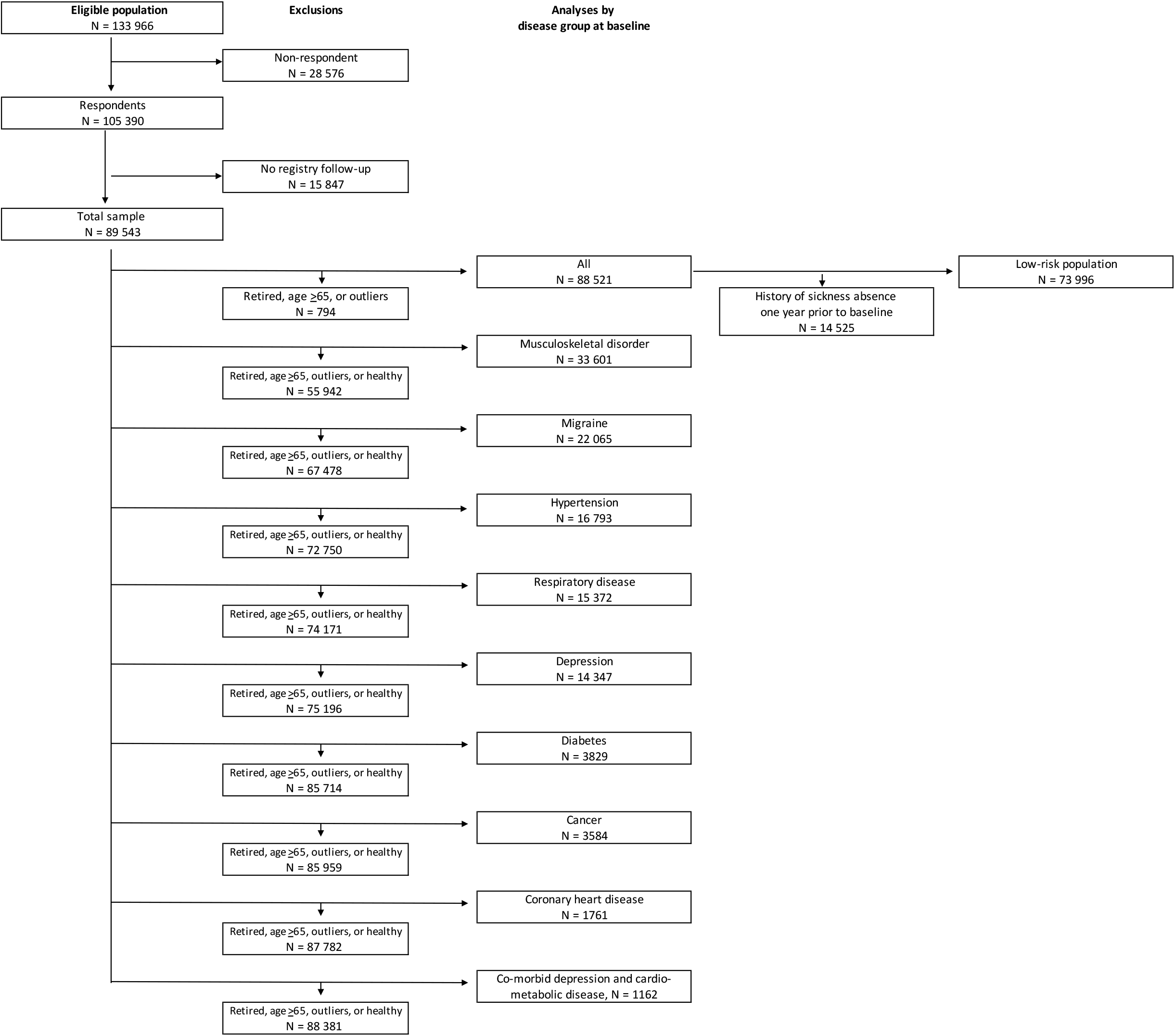
Cause of work disability at follow-up in all participants and subgroups

The unadjusted bivariate associations between 105 predictor variables and work disability are illustrated in **figure 3**. As expected, all variables from the existing 8-item risk score (age, self-rated health, number of sickness absences in previous year, socioeconomic position, chronic illnesses, sleep problems, body mass index, and smoking) were strongly associated with work disability. Many other health-related variables were also strongly associated with work disability whereas the associations of the remaining items related to risk behaviours, work characteristics, team climate and management were weaker. The pattern of findings was similar in all disease groups (**appendix**, p. 7-15).

**Figure 3.**
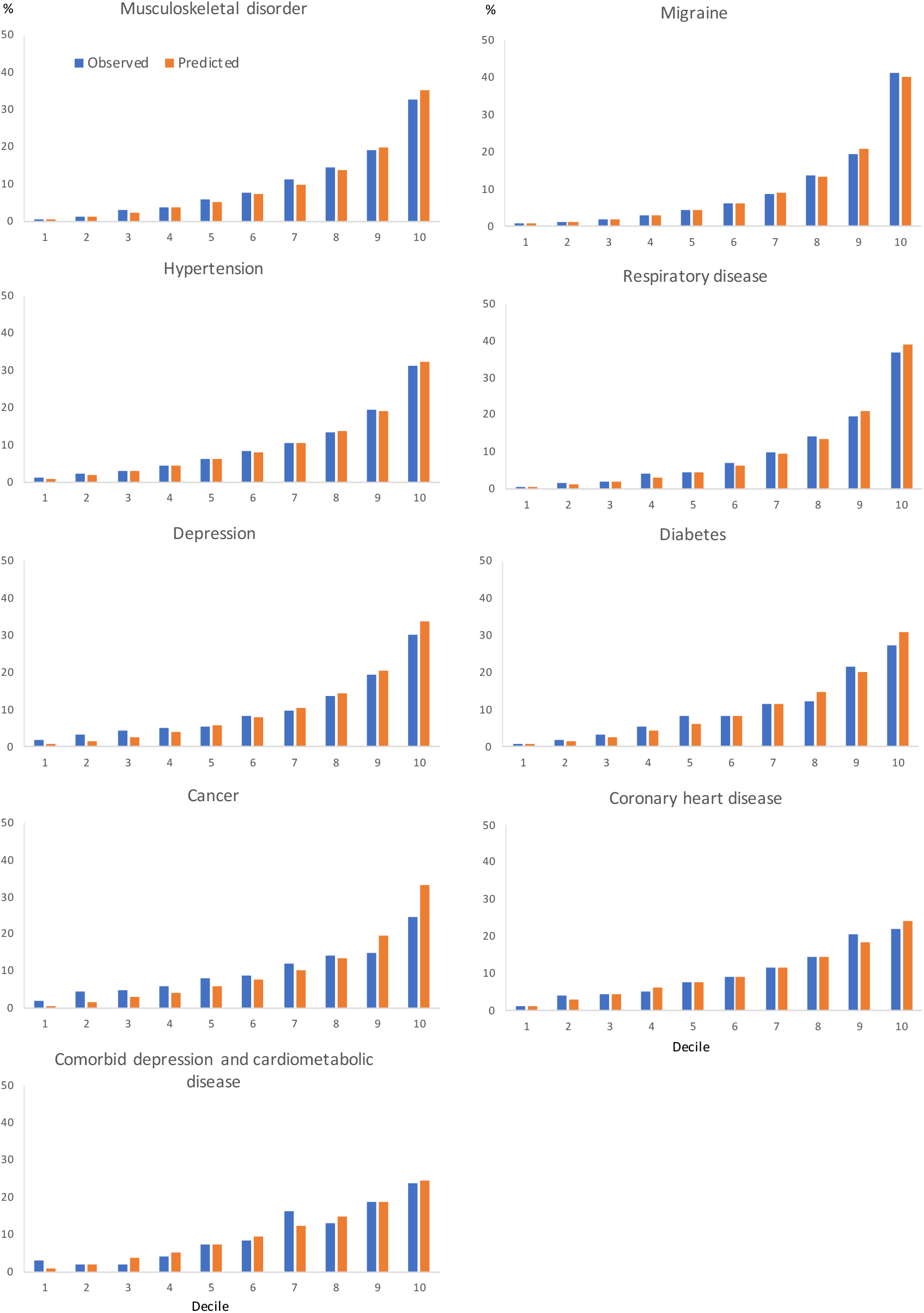
Bivariate association between individual predictor items at baseline and work disability at follow-up in all participants with and without chronic disease at baseline

To examine whether the existing model could be improved, we developed two alternative models in each disease group (analysis of re-estimated coefficients and new predictive algorithms is provided in **appendix** p. 16-17.) The predictors in the re-estimated models included between 4 and 7 of the 8 predictors in the existing model. Each of these models included age, socioeconomic status, self-rated health and the number of sickness absences in previous year. Individual self-reported chronic diseases were also frequently part of the models. Some work-related items were also included in some of the disease groups.

**Table 2** shows that C-statistics was similar as in the original study[4] for all employees with and without chronic conditions (0.84, 95% CI: 0.84 to 0.85) and among those with no sickness absence at baseline (0.82, 95% CI: 0.81 to 0.83). The table also provides a comparison of the performance between the existing model and the two alternatives. The existing model performed well in all disease groups. The C-statistics was ≥0.80 in those with musculoskeletal disorders (0.80, 95% CI: 0.80 to 0.81), migraine (0.83, 95% CI: 0.82 to 0.84) and respiratory disease (0.82, 95% CI: 0.81 to 0.83). For all other subgroups, including hypertension, depression, diabetes, cancer, coronary heart disease or comorbid depression and cardiometabolic disease, C-statistics was ≥0.72 but less than 0.80. The C-statistics for the re-estimated model and the new model were virtually the same as for the existing model, suggesting no improvement in predictive performance. Calibration plots for the existing model indicated a high correspondence between the predicted and the observed risk in all disease groups (**figure 4**).

**Table 2.**
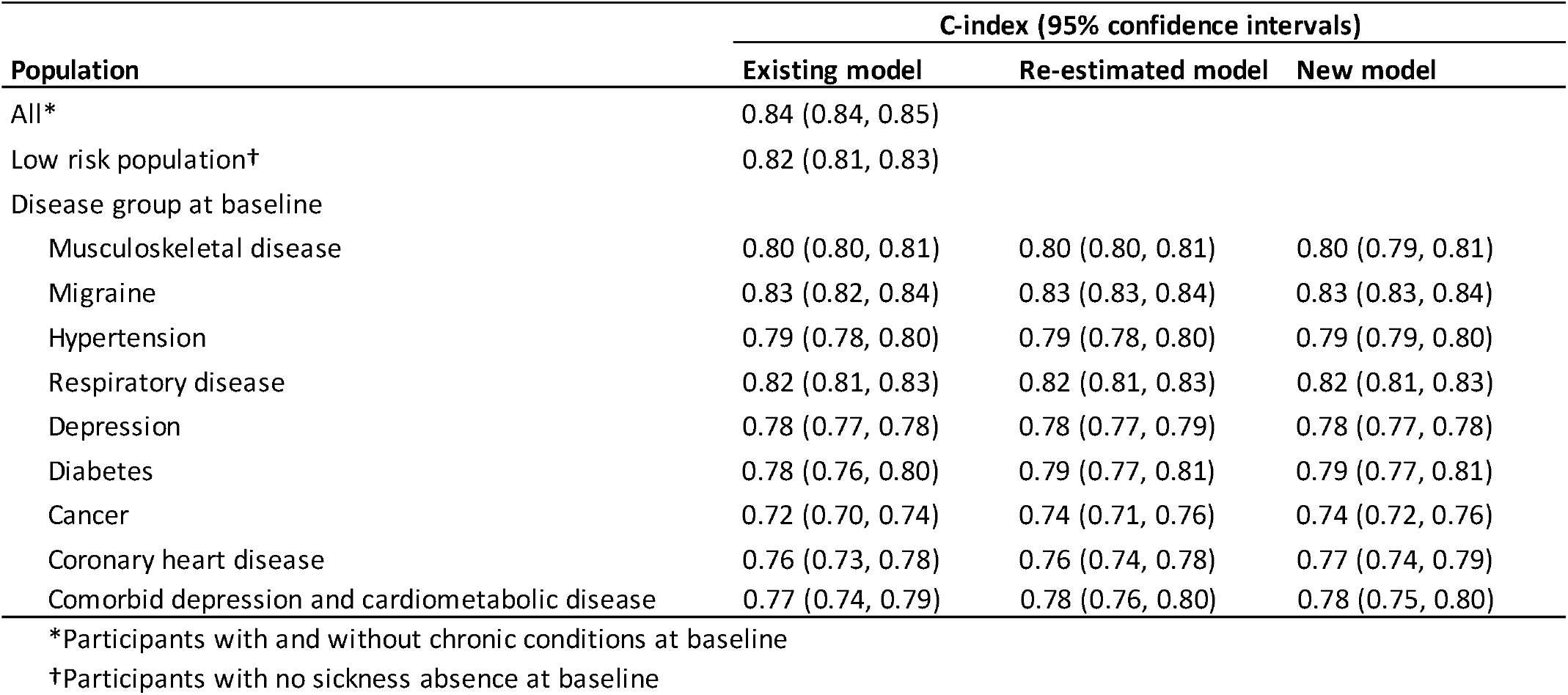
C-index for the existing, re-estimated and new prediction models in all participants and subgroups of individuals with no history of sickness absence and those with a chronic condition at baseline

| Population | C-index (95% confidence intervals) |  |  |
| --- | --- | --- | --- |
|  | Existing model | Re-estimated model | New model |
| All* | 0.84 (0.84, 0.85) |  |  |
| Low risk population† | 0.82 (0.81, 0.83) |  |  |
| Disease group at baseline |  |  |  |
| Musculoskeletal disease | 0.80 (0.80, 0.81) | 0.80 (0.80, 0.81) | 0.80 (0.79, 0.81) |
| Migraine | 0.83 (0.82, 0.84) | 0.83 (0.83, 0.84) | 0.83 (0.83, 0.84) |
| Hypertension | 0.79 (0.78, 0.80) | 0.79 (0.78, 0.80) | 0.79 (0.79, 0.80) |
| Respiratory disease | 0.82 (0.81, 0.83) | 0.82 (0.81, 0.83) | 0.82 (0.81, 0.83) |
| Depression | 0.78 (0.77, 0.78) | 0.78 (0.77, 0.79) | 0.78 (0.77, 0.78) |
| Diabetes | 0.78 (0.76, 0.80) | 0.79 (0.77, 0.81) | 0.79 (0.77, 0.81) |
| Cancer | 0.72 (0.70, 0.74) | 0.74 (0.71, 0.76) | 0.74 (0.72, 0.76) |
| Coronary heart disease | 0.76 (0.73, 0.78) | 0.76 (0.74, 0.78) | 0.77 (0.74, 0.79) |
| Comorbid depression and cardiometabolic disease | 0.77 (0.74, 0.79) | 0.78 (0.76, 0.80) | 0.78 (0.75, 0.80) |

\*Participants with and without chronic conditions at baseline
†Participants with no sickness absence at baseline

**Figure 4.**
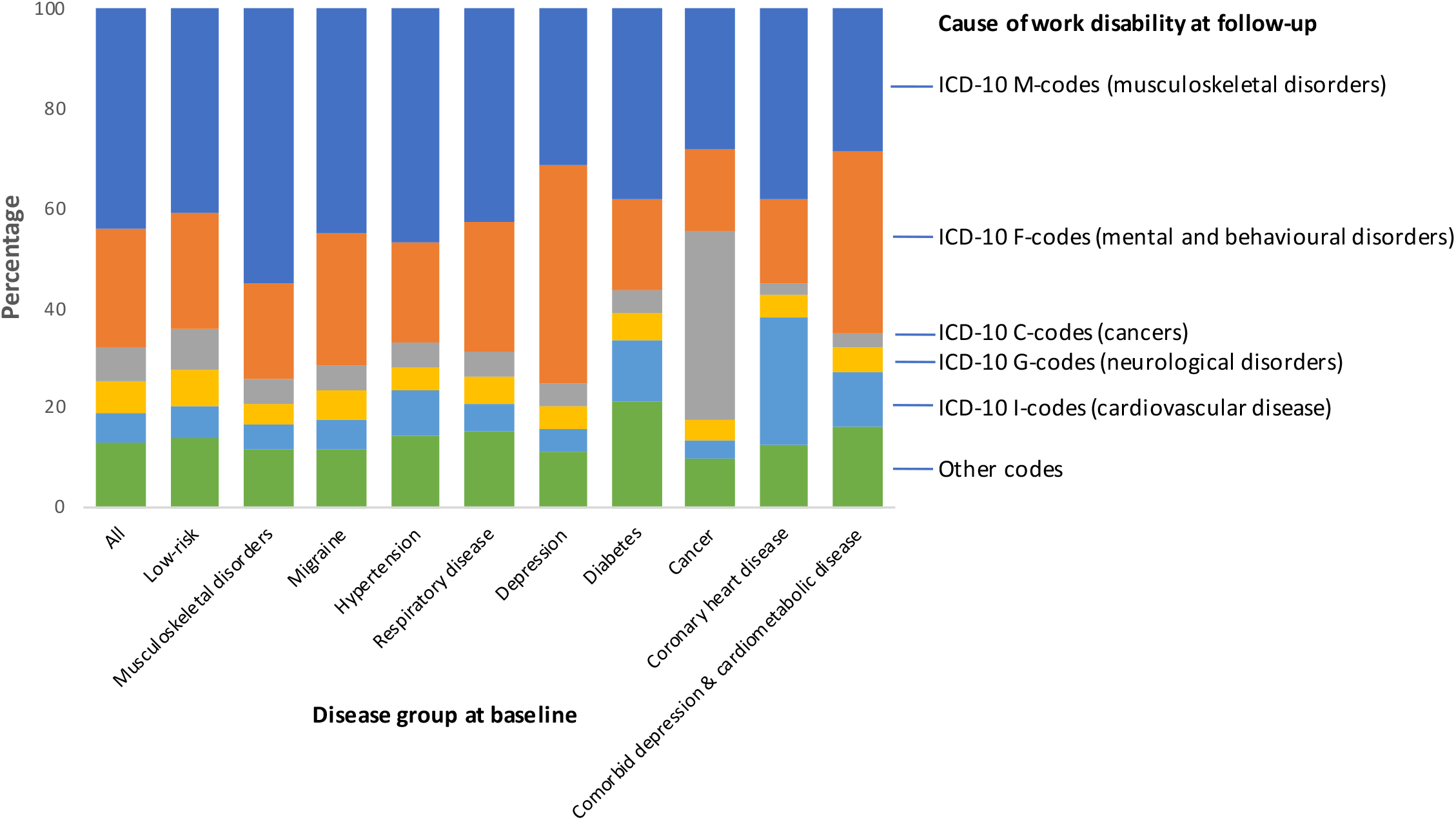
Observed and predicted incidence of work disability by deciles of the work disability risk score

**Table 3** shows the detection and false positive rates for dichotomized existing risk scores using various risk thresholds (for sensitivity, specificity and positive and negative predictive value, see **appendix**, p. 18). With a high threshold (≥50% predicted probability indicating high risk), the false positive rate ranged between 2.6% and 8.6% in all disease groups. Exceptions were for participants with coronary heart disease (19.0%) and those with comorbid depression and cardiometabolic disease (20.1%) where it was markedly higher. The detection rate varied between 18.9% (participants with cancer) and 52.8% (participants with comorbid depression and cardiometabolic disease) and the ratio of true to false positives was 1 : 1.0 to 1.4. With a low threshold (5% predicted probability indicating high risk), detection rate raised to 92.0% to 99.2% (less than 1 in 10 disability cases were missed), but with high false positive rate (54.2% to 94.4% depending on the disease group). For both the 50% and 5% thresholds, the detection and false positive rates were slightly lower in the total working population, but the ratio of true to false positives was approximately the same as in the population with chronic diseases.

**Table 3.** Detection rate, false positive rate and the ratio true-to-false positives for the existing prediction model in all participants and subgroups of individuals with no history of sickness absence and those with a chronic condition at baseline

| Predictive performance for a positive test | Cut-off (%) for a positive test result |  |  |  |  |  |  |
| --- | --- | --- | --- | --- | --- | --- | --- |
|  | 5 | 10 | 20 | 30 | 40 | 50 | 60 |
| **Musculoskeletal disorders** |  |  |  |  |  |  |  |
| Detection rate | 95.3 | 86.6 | 65.9 | 46.6 | 32.7 | 21.1 | 12.8 |
| False positive rate | 70.2 | 50.4 | 26.6 | 13.6 | 7.1 | 3.6 | 1.7 |
| Ratio true to false positives | 1 : 4.5 | 1 : 3.5 | 1 : 2.5 | 1 : 1.8 | 1 : 1.3 | 1 : 1.0 | 1 : 0.8 |
| **Migraine** |  |  |  |  |  |  |  |
| Detection rate | 92.0 | 83.1 | 63.6 | 47.6 | 33.3 | 22.1 | 14.3 |
| False positive rate | 54.2 | 37.0 | 19.3 | 10.0 | 5.2 | 2.6 | 1.2 |
| Ratio true to false positives | 1 : 5.4 | 1 : 4.1 | 1 : 2.8 | 1 : 1.9 | 1 : 1.4 | 1 : 1.1 | 1 : 0.7 |
| **Hypertension** |  |  |  |  |  |  |  |
| Detection rate | 96.4 | 89.6 | 71.2 | 53.2 | 37.3 | 24.8 | 15.2 |
| False positive rate | 80.5 | 61.7 | 34.4 | 17.9 | 9.4 | 4.9 | 2.2 |
| Ratio true to false positives | 1 : 4.7 | 1 : 3.9 | 1 : 2.7 | 1 : 1.9 | 1 : 1.4 | 1 : 1.1 | 1 : 0.8 |
| **Respiratory disease** |  |  |  |  |  |  |  |
| Detection rate | 93.0 | 84.8 | 66.0 | 49.9 | 35.8 | 24.1 | 15.1 |
| False positive rate | 59.9 | 42.9 | 23.2 | 12.9 | 7.2 | 3.8 | 1.8 |
| Ratio true to false positives | 1 : 4.6 | 1 : 3.6 | 1 : 2.5 | 1 : 1.9 | 1 : 1.4 | 1 : 1.1 | 1 : 0.8 |
| **Depression** |  |  |  |  |  |  |  |
| Detection rate | 93.4 | 86.2 | 69.7 | 53.7 | 40.1 | 27.4 | 18.0 |
| False positive rate | 72.3 | 54.3 | 31.7 | 18.5 | 10.7 | 6.1 | 3.0 |
| Ratio true to false positives | 1 : 3.9 | 1 : 3.2 | 1 : 2.3 | 1 : 1.7 | 1 : 1.3 | 1 : 1.1 | 1 : 0.8 |
| **Diabetes** |  |  |  |  |  |  |  |
| Detection rate | 97.0 | 91.6 | 77.7 | 62.4 | 46.3 | 32.8 | 20.7 |
| False positive rate | 80.8 | 68.0 | 46.3 | 27.6 | 16.1 | 8.6 | 4.3 |
| Ratio true to false positives | 1 : 4.2 | 1 : 3.8 | 1 : 3.0 | 1 : 2.3 | 1 : 1.8 | 1 : 1.3 | 1 : 1.1 |
| **Cancer** |  |  |  |  |  |  |  |
| Detection rate | 93.1 | 80.4 | 58.3 | 42.2 | 30.3 | 18.9 | 13.2 |
| False positive rate | 78.8 | 59.3 | 33.5 | 18.8 | 9.7 | 4.3 | 2.1 |
| Ratio true to false positives | 1 : 5.1 | 1 : 4.5 | 1 : 3.5 | 1 : 2.7 | 1 : 1.9 | 1 : 1.4 | 1 : 1.0 |
| **Coronary heart disease** |  |  |  |  |  |  |  |
| Detection rate | 99.2 | 97.2 | 88.9 | 76.1 | 62.8 | 47.2 | 31.7 |
| False positive rate | 94.4 | 87.5 | 66.4 | 45.5 | 30.3 | 19.0 | 10.3 |
| Ratio true to false positives | 1 : 3.3 | 1 : 3.1 | 1 : 2.6 | 1 : 2.0 | 1 : 1.7 | 1 : 1.4 | 1 : 1.1 |
| **Comorbid depression and cardiometabolic disease** |  |  |  |  |  |  |  |
| Detection rate | 98.1 | 96.3 | 90.7 | 79.8 | 67.7 | 52.8 | 38.5 |
| False positive rate | 89.0 | 79.9 | 61.9 | 45.2 | 32.3 | 20.1 | 11.8 |
| Ratio true to false positives | 1 : 2.4 | 1 : 2.2 | 1 : 1.8 | 1 : 1.5 | 1 : 1.2 | 1 : 1.0 | 1 : 0.8 |
| **Low-risk population** |  |  |  |  |  |  |  |
| Detection rate | 81.6 | 63.5 | 35.0 | 17.2 | 8.3 | 3.5 | 1.2 |
| False positive rate | 38.2 | 21.5 | 7.7 | 2.7 | 0.9 | 0.4 | 0.1 |
| Ratio true to false positives | 1 : 8.1 | 1 : 5.9 | 1 : 3.8 | 1 : 2.7 | 1 : 2.0 | 1 : 1.9 | 1 : 1.6 |
| **Total cohort** |  |  |  |  |  |  |  |
| Detection rate | 87.9 | 75.0 | 52.5 | 35.9 | 24.0 | 15.0 | 9.1 |
| False positive rate | 43.7 | 27.0 | 12.0 | 5.7 | 2.8 | 1.4 | 0.62 |
| Ratio true to false positives | 1 : 5.9 | 1 : 4.3 | 1 : 2.7 | 1 : 1.9 | 1 : 1.4 | 1 : 1.1 | 1 : 0.8 |

## Discussion

Our study shows that a short, self-administered survey instrument has predictive utility for work disability in people with chronic conditions and comorbidity. This survey-based risk calculator includes age, self-rated health, number of sickness absences, socioeconomic position, the number of comorbidities, sleep problems, body mass index, and smoking habit. This algorithm can be used in many settings, including members of the public who have web access, and the assessment does not require laboratory testing or other clinical measurements. The calculator might be used to identify working-age people with common chronic diseases with an elevated risk of future work disability and so facilitate early intervention.

Approximately one third of people at age 40 have a chronic condition and the proportion rises to 75% by age 65.[19] Despite this high prevalence and the urgent need for new measures to prevent their work disability, we are not aware of other large-scale studies on risk stratification algorithms for work disability in employees with chronic conditions. In our study, C-index exceeded 0.72 in all disease groups and was 0.80 or greater for those with musculoskeletal disorders, migraine and respiratory disease. These results indicate good discrimination and are comparable to those reported for established risk prediction tool currently used in clinical practice. For example, the C-index is 0.72 for the Pooled Cohort Equations to predict the 10-year risk of cardiovascular disease events using 9 risk factors (age, sex, race, diabetes status, smoking status, antihypertensive medication use, total cholesterol, HDL cholesterol levels, and systolic blood pressure);[20] between 0.74 and 0.77 for the FINDRISC model to predict the risk of type 2 diabetes using 8 characteristics (age, sex, BMI, antihypertensive medication use, blood glucose, physical activity, diet and family history);[21, 22] and 0.70 to 0.91 for QRISK3 to predict future cardiovascular disease based on a wide range of risk factors obtained from electronic health records (age, sex, ethnicity, socioeconomic deprivation, angina or heart attack in a 1st degree relative at age < 60, chronic kidney disease, migraine, corticosteroids, systemic lupus erythematosus, use of atypical antipsychotics, severe mental illness, steroid treatment, erectile dysfunction, total/ HDL cholesterol ratio, BMI, systolic blood pressure variability).[23]

In clinical decision making, dichotomised predictive scores are used to distinguish patients who should receive intervention or referrals for further assessments, although few studies have reported relevant performance metrics in this regard.[12] While the performance of our dichotomized work disability risk score fell short of the best established clinical screening tests, such as mammography for breast cancer (detection rate 75% with a false positive rate of 8%)[24, 25] and faecal immunochemical test (FIT) for colon cancer (79%/6%),[26] it was similar to those reported for widely used cardiovascular disease risk scores, such as QRISK2 (detection and false positive rates 40% and 13% for men and 26% and 6% for women)[12] and thus appears to provide a useful tool to aid decisions of targeting preventive interventions. More specifically, our risk calculator had a relatively high false positive rate for test positive thresholds that allowed high detection rates. Conversely, the use of a threshold that provided low (approximately 5%) false positive rate, resulted in detection of only 20-25% of disability cases. This means that the score is useful in informing the targeting of interventions with no significant harm from overtreatment and in informing referrals to more detailed assessments for potential tailored preventive measures. By contrast, the score should not be used for expensive or new interventions with uncertain safety profiles as many people who will not benefit from the intervention will be targeted.

The present study benefits from the use of data which are from a country (Finland) where ascertainment of work disability pension was possible with linkage to comprehensive records from the national pension register with virtually full coverage for those in employment. However, this study has several limitations. Although work disability is defined by impairment, receipt of a disability pension is additionally dependent on non-medical factors, such as disability pension regulations, the work environment, the nature of the job, and the extent to which a workplace is prepared to accommodate the disability. The generalizability of the present findings should therefore be investigated in other countries with different disability pension policies. Our study largely comprised women and all study participants were drawn from public sector workplaces. Replications in different study populations and settings are therefore required.

In conclusion, detection of individuals at high risk is a precondition for effective targeted interventions to prevent long-term work disability and a basis for developing cost-effective strategies to avoid early labour-market exit. Predictive performance of a simple, widely applicable, cost-free work disability risk score was comparable to those observed for established risk scores for other outcomes. These findings suggest that it is possible to predict work disability in a working population with chronic disease using a scalable internet-based tool with a reasonable accuracy and thus aid decisions of targeted interventions and referrals to detailed assessments of tailored interventions.

## Supporting information

STROBE-checklist

## Data Availability

Pseudonymised questionnaire data from the FPS study can be shared upon request to the investigators. Linked health records require separate permission from the Findata, the Health and Social Data Permit Authority in Finland.

## Statements and Declarations

### Author contributions

All authors participated in designing the study, generating hypotheses, interpreting the data, and critically reviewing the paper. STN, with MK, wrote the first draft of the report. STN, with help from JA and JP, performed data analysis. STN, JA, JP, and MK had full access to all the data. The corresponding author attests that all listed authors meet authorship criteria and that no others meeting the criteria have been omitted.

### Ethics approval

Ethical approval was obtained from the ethics committee of the Helsinki-Uusimaa Hospital District Ethics Committee (HUS/1210/2016).

### Consent to participate

Informed consent was obtained from all individual participants included in the study.

### Data sharing

Statistical syntax for the analysis of the present study is available in Appendix, pp. 19–23. Pseudonymised questionnaire data from the FPS study can be shared upon request to the investigators. Linked health records require separate permission from the Findata, the Health and Social Data Permit Authority in Finland.

