## Appendix for "Predicting work disability among people with chronic conditions: a large prospective cohort study"

Contents Page


*Table S1*. Potential predictors 2

*Table S2*. Coefficients for formulating linear predictor in the existing model 5

*Table S3*. Cause of work disability at follow-up in all participants and subgroups 6

*Figure S1.* Bivariate association between individual predictor items at baseline and work disability at follow-up in participants with musculoskeletal disorder at baseline 7

*Figure S2.* Bivariate association between individual predictor items at baseline and work disability at follow-up in participants with migraine at baseline 8

*Figure S3.* Bivariate association between individual predictor items at baseline and work disability at follow-up in participants with hypertension at baseline 9

*Figure S4.* Bivariate association between individual predictor items at baseline and work disability at follow-up in participants with respiratory disease at baseline 10

*Figure S5.* Bivariate association between individual predictor items at baseline and work disability at follow-up in participants with depression at baseline 11

*Figure S6.* Bivariate association between individual predictor items at baseline and work disability at follow-up in participants with diabetes at baseline 12

*Figure S7.* Bivariate association between individual predictor items at baseline and work disability at follow-up in participants with cancer at baseline 13

*Figure S8.* Bivariate association between individual predictor items at baseline and work disability at follow-up in participants with coronary heart disease at baseline 14

*Figure S9.* Bivariate association between individual predictor items at baseline and work disability at follow-up in participants with comorbid depression and cardiometabolic disease (diabetes, CHD or stroke) at baseline 15

*Table S4*. Re-estimated coefficients and new predictive algorithms 16

*Table S5.* Sensitivity, specificity and positive and negative predictive value for the existing prediction model in all participants and subgroups of individuals with no history of sickness absence and those with a chronic condition at baseline 18

Statistical code 19

**Table S1. Potential predictors**

| **Variable** |
|---|
| Age |
| Sex |
| Socioeconomic status |
| BMI |
| Height in cm |
| Weight in kg |
| Smoking |
| Do you smoke or have you smoked regularly (every day or almost every day)? |
| Do you still smoke regularly? |
| Alcohol consumption |
| Have you ever had at least a glass of an alcoholic beverage? |
| How many times a week you consume beer? |
| …wine? |
| …spirits? |
| How many times have you passed out from drinking during the past year? |
| Inactivity |
| During the past year, how many hours in a week have you walked? |
| …walked briskly? |
| …jogged? |
| …ran? |
| GHQ |
| In past weeks have you been able to concentrate? |
| ...loss of sleep over worry |
| ...playing a useful part |
| ...capable of making decisions |
| ...felt constantly under strain |
| ...couldn’t overcome difficulties |
| ...able to enjoy day-to-day activities |
| ...able to face problems |
| ...feeling unhappy and depressed |
| ...losing confidence |
| ...thinking of self as worthless |
| ...feeling reasonably happy |
| Number of chronic diseases |
| Allergic rhinitis |
| Asthma or chronic bronchitis |
| Hypertension |
| Diabetes |
| Myocardial infarction or angina pectoris |
| Cerebrovascular disorder (transient ischaemic attack) |
| Gastric or duodenal ulcer |
| Migraine |
| Depression |
| Other mental disorder |

Cancer
Musculoskeletal disorder (i.e. arthrosis, rheumatoid arthritis or sciatica)
Self-rated health
Jenkins sleep scale
- How many times in the past 4 weeks have you had ...trouble falling a sleep
- …frequent awakenings during the night
- ...trouble remaining asleep
- ...feelings of fatigue and sleepiness despite receiving a typical night's rest

No. of sickness absences in previous year
Job strain
Job control
- My work requires creativity
- My work requires me to learn new things
- My work involves a lot of repetitive tasks
- I have a say in the tasks included in my work
- My work requires highly developed skills
- I have very little freedom to decide how I do my work

Job demand
- My work requires a lot of effort
- I am expected to do unreasonable amount of work
- I have sufficient time to get my work done

Relational justice
- Your supervisor considers your viewpoint
- Your supervisor is able to suppress personal biases
- Your supervisor provides you with timely feedback about the decision and its implications
- Your supervisor treats you with kindness and consideration
- Your supervisor shows concern for your rights as an employee
- Your supervisor takes steps to deal with you in a truthful manner

Procedural justice
- Procedures designed to… collect accurate information necessary for making decisions
- …provide opportunities to appeal or challenge the decision
- …have all sides affected by the decision represented.
- …generate standards so that decision could be made with consistency.
- …hear the concerns of all those affected by the decision.
- …provide useful feedback regarding the decision and its implementation
- …allow for requests for clarification or additional information about the decision.

Participatory safety
- People keep each other informed about work-related issues in the team
- There are real attempts to share information throughout the team
- We have a "we are in it together" attitude
- People feel understood and accepted by each other

Support for innovation
- People in this team are always searching for fresh, new ways of looking at problems
- In this team we take the time needed to develop new ideas
- People in the team co-operate in order to help develop and apply new ideas

Vision

To what extent do you think your team's objectives are clearly understood by other members of the team?

How far are you in agreement with these objectives?

To what extent do you think your team's objectives can actually be achieved?

How worthwhile do you think these objectives are?

Task orientation

Are team members prepared to question the basis of what the team is doing?

Does the team critically appraise potential weaknesses in what it is doing in order to achieve the best possible outcome?

Do members of the team build on each other's ideas in order to achieve the best possible outcome?

Social capital at work place

Do members of the team build on each other's ideas in order to achieve the best possible outcome?

People keep each other informed about work-related issues in the team

We have a "we are in it together" attitude

People feel understood and accepted by each other

People in the team co-operate in order to help develop and apply new ideas

Do members of the team build on each other's ideas in order to achieve the best possible outcome?

Your supervisor treats you with kindness and consideration

Your supervisor shows concern for your rights as an employee

Your supervisor takes steps to deal with you in a truthful manner

Effort-Reward imbalance

Effort

How much of your skills and resources you invest in your work?

Reward

Do you feel that you get value for money for your work?

Do you feel that you get recognition and respect for your work?

Do you feel that you get personal satisfaction of your work?

Shift work

Night shift

**Table S2. Coefficients for formulating linear predictor in the existing model**

| Predictor | Beta coefficient |
|---|---|
| Age <35 | 0 |
| Age=35-39 | -0.2339 |
| Age=40-44 | -0.4356 |
| Age=45-49 | -0.8825 |
| Age=50-54 | -1.2873 |
| Age=55+ | -1.5418 |
| BMI<18.5 | -0.1724 |
| BMI 18.5-24.9 | 0 |
| BMI=25-29.9 | -0.0668 |
| BMI=30+ | -0.1753 |
| SEP=1 | 0 |
| SEP=2 | -0.0457 |
| SEP=3 | -0.3171 |
| SEP=4 | -0.3213 |
| SEP=5 | -0.546 |
| SEP=6 | -0.5294 |
| SEP=7 | -0.6597 |
| Smoking=NO | 0 |
| Smoking=YES | -0.1638 |
| Chronic illness=0 | 0 |
| Chronic illness=1 | -0.2252 |
| Chronic illness=2 | -0.4462 |
| Chronic illness=3 | -0.5342 |
| Self-rated health=1 | 0 |
| Self-rated health=2 | -0.2348 |
| Self-rated health=3 | -0.5539 |
| Self-rated health=4 | -1.1336 |
| Self-rated health=5 | -1.5182 |
| Difficulty falling asleep=1 | 0 |
| Difficulty falling asleep=2 | -0.0281 |
| Difficulty falling asleep=3 | -0.0769 |
| Difficulty falling asleep=4 | -0.1267 |
| Difficulty falling asleep=5 | -0.2014 |
| Difficulty falling asleep=6 | -0.2245 |
| No. sickness absences in previous year=0 | 0 |
| No. sickness absences in previous year=1 | -0.4334 |
| No. sickness absences in previous year=2 | -0.7413 |
| No. sickness absences in previous year=3 | -1.133 |

Intercept = 5.7912

Scale = 1.2046

### Table S3. Cause of work disability at follow-up in all participants and subgroups

| | All | Low-risk population* | Disease group at baseline | | | | | | | | |
|---|---|---|---|---|---|---|---|---|---|---|---|
| | | | MSD | Migraine | Hypertension | Respiratory disease | Depression | Diabetes | Cancer | CHD | DCD |
| | 88521 | 73996 | 33601 | 22065 | 16793 | 15372 | 14347 | 3829 | 3584 | 1761 | 1162 |
| Mean age (SD) at baseline | 43.1 (9.8) | 42.7 (9.9) | 48.5 (8.4) | 43.4 (9.7) | 50.8 (7.5) | 45.1 (9.8) | 45.9 (9) | 49.5 (9.3) | 50.8 (8) | 53.5 (7) | 50.2 (8.4) |
| Mean follow-up time | 8.6 | 8.8 | 7.5 | 8.4 | 7.1 | 7.9 | 7.8 | 6.8 | 6.7 | 5.9 | 6.3 |
| N of work disability cases at follow-up | 6836 | 4029 | 4734 | 2186 | 2539 | 1883 | 2392 | 628 | 509 | 398 | 322 |
| Insidence per 1000 person-years | 8.9 | 6.2 | 18.7 | 11.8 | 21.3 | 15.5 | 21.5 | 24.3 | 21.1 | 38.4 | 44.3 |
| Cause of disability pension, N (%) | | | | | | | | | | | |
| ICD-10 M-codes (musculoskeletal disorders) | 3023 (44.2) | 1654 (41.0) | 2608 (55.1) | 982 (44.9) | 1191 (46.9) | 812 (43.1) | 749 (31.3) | 240 (38.2) | 144 (28.3) | 154 (38.7) | 93 (28.9) |
| ICD-10 F-codes (mental and behavioural disorders) | 1618 (23.7) | 930 (23.1) | 892 (18.8) | 576 (26.3) | 511 (20.1) | 482 (25.6) | 1049 (43.9) | 114 (18.2) | 82 (16.1) | 67 (16.8) | 116 (36) |
| ICD-10 C-codes (cancers) | 472 (6.9) | 331 (8.2) | 243 (5.1) | 118 (5.4) | 124 (4.9) | 97 (5.2) | 111 (4.6) | 28 (4.5) | 193 (37.9) | 9 (2.3) | 10 (3.1) |
| ICD-10 G-codes (neurological disorders) | 425 (6.2) | 294 (7.3) | 210 (4.4) | 129 (5.9) | 117 (4.6) | 99 (5.3) | 109 (4.6) | 36 (5.7) | 20 (3.9) | 17 (4.3) | 15 (4.7) |
| ICD-10 I-codes (cardiovascular disease) | 398 (5.8) | 259 (6.4) | 229 (4.8) | 123 (5.6) | 225 (8.9) | 103 (5.5) | 102 (4.3) | 76 (12.1) | 19 (3.7) | 102 (25.6) | 36 (11.2) |
| Other codes | 900 (13.2) | 562 (13.9) | 552 (11.7) | 258 (11.8) | 371 (14.6) | 290 (15.4) | 272 (11.4) | 134 (21.3) | 51 (10) | 49 (12.3) | 52 (16.1) |

*Abbreviations* : CHD, coronary heart disease; DCD, comorbid depression and cardiometabolic disease (diabetes or CHD); MSD, musculoskeletal disorder

*Participants with no sickness absence during one year before baseline

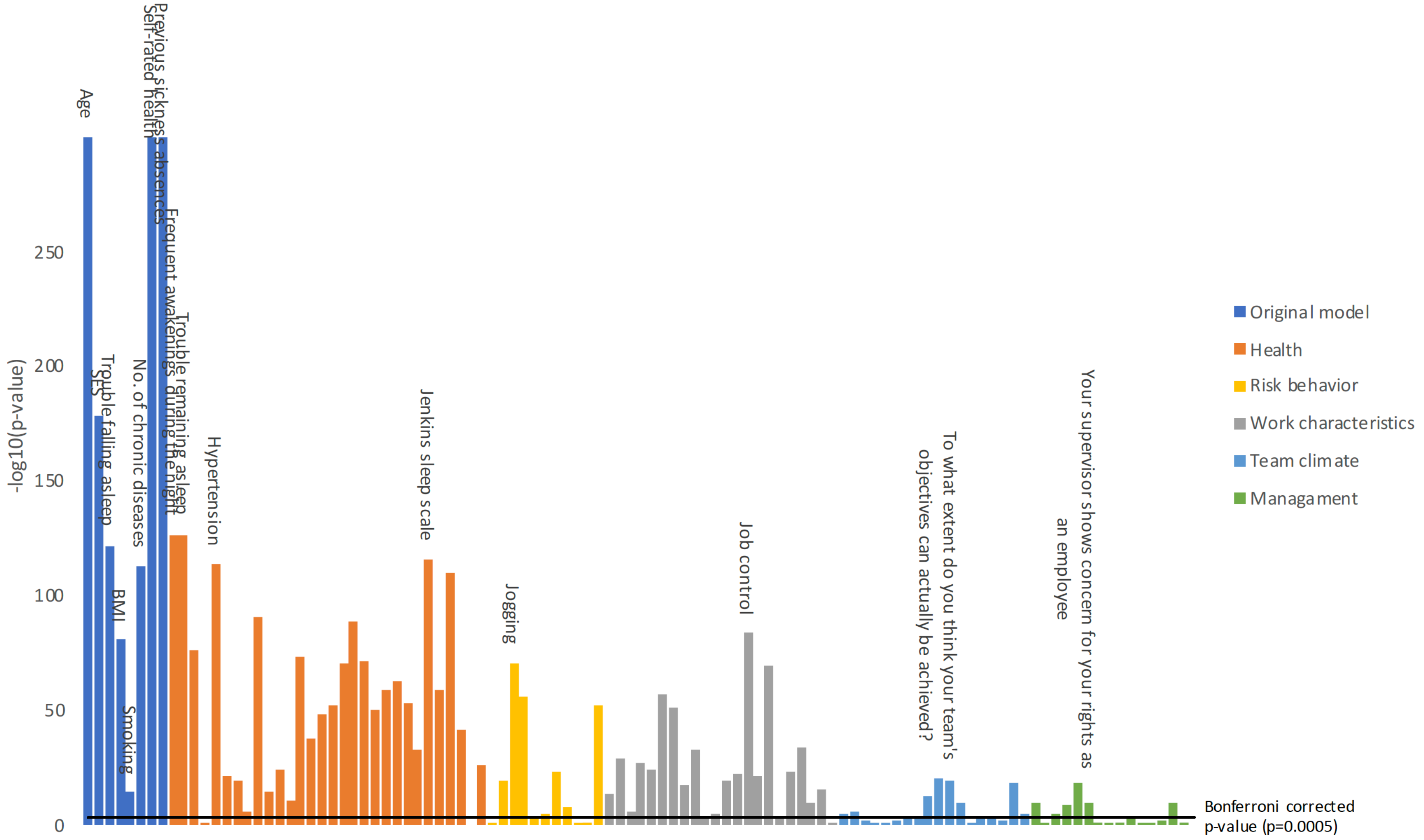


**Figure S1. Bivariate association between individual predictor items at baseline and work disability at follow-up in participants with musculoskeletal disorder at baseline**

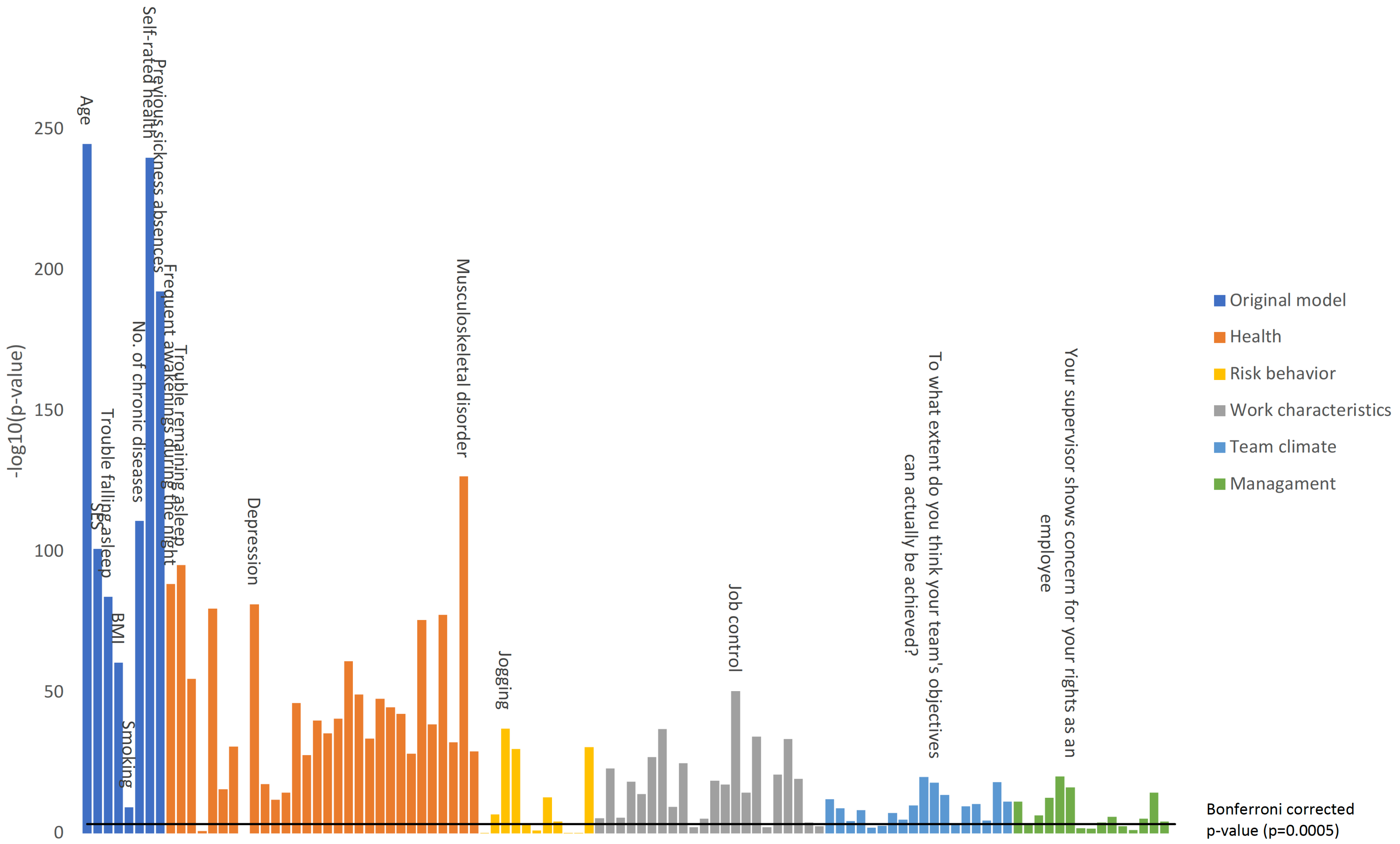


**Figure S2. Bivariate association between individual predictor items at baseline and work disability at follow-up in participants with migraine at baseline**

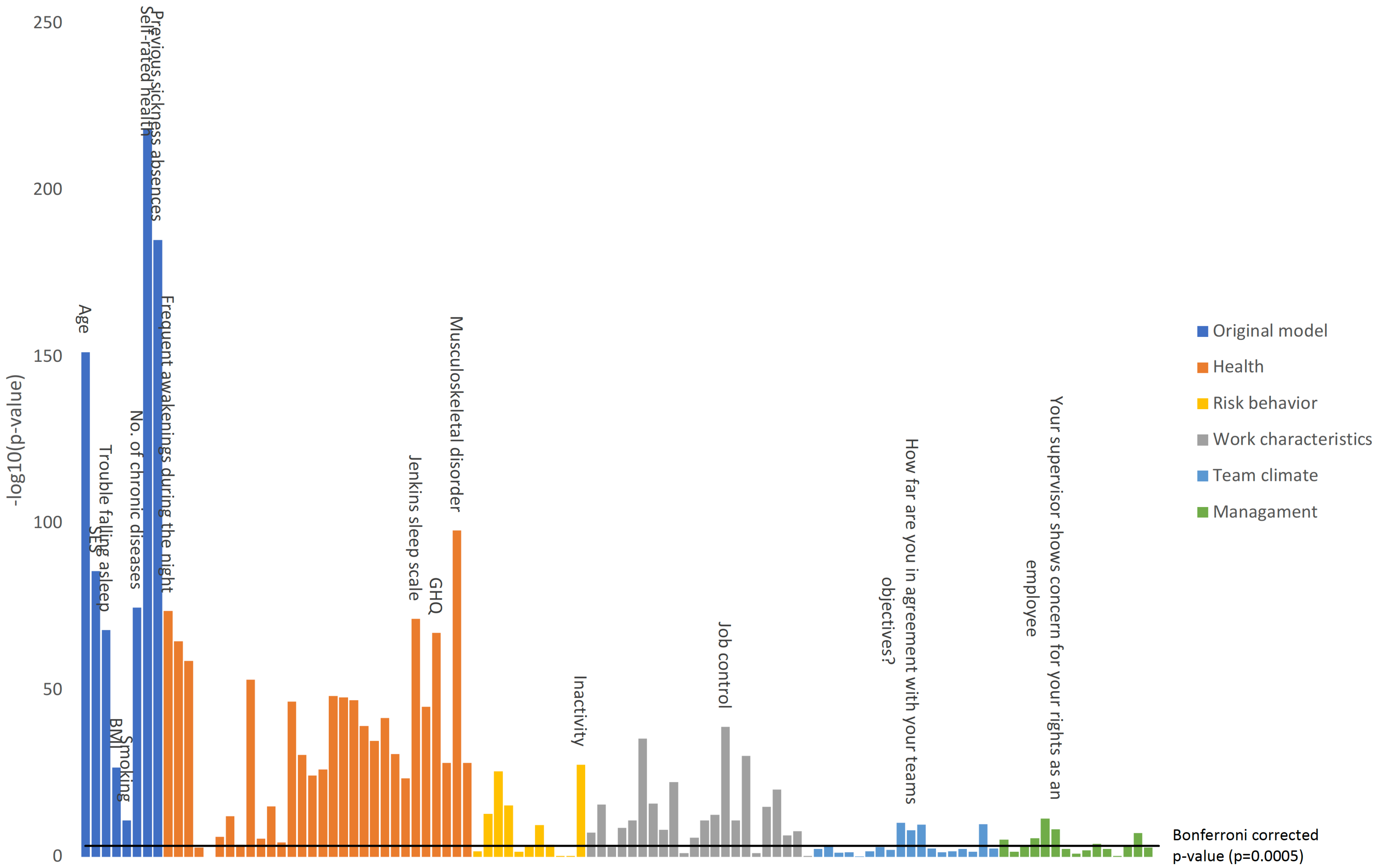


**Figure S3. Bivariate association between individual predictor items at baseline and work disability at follow-up in participants with hypertension at baseline**

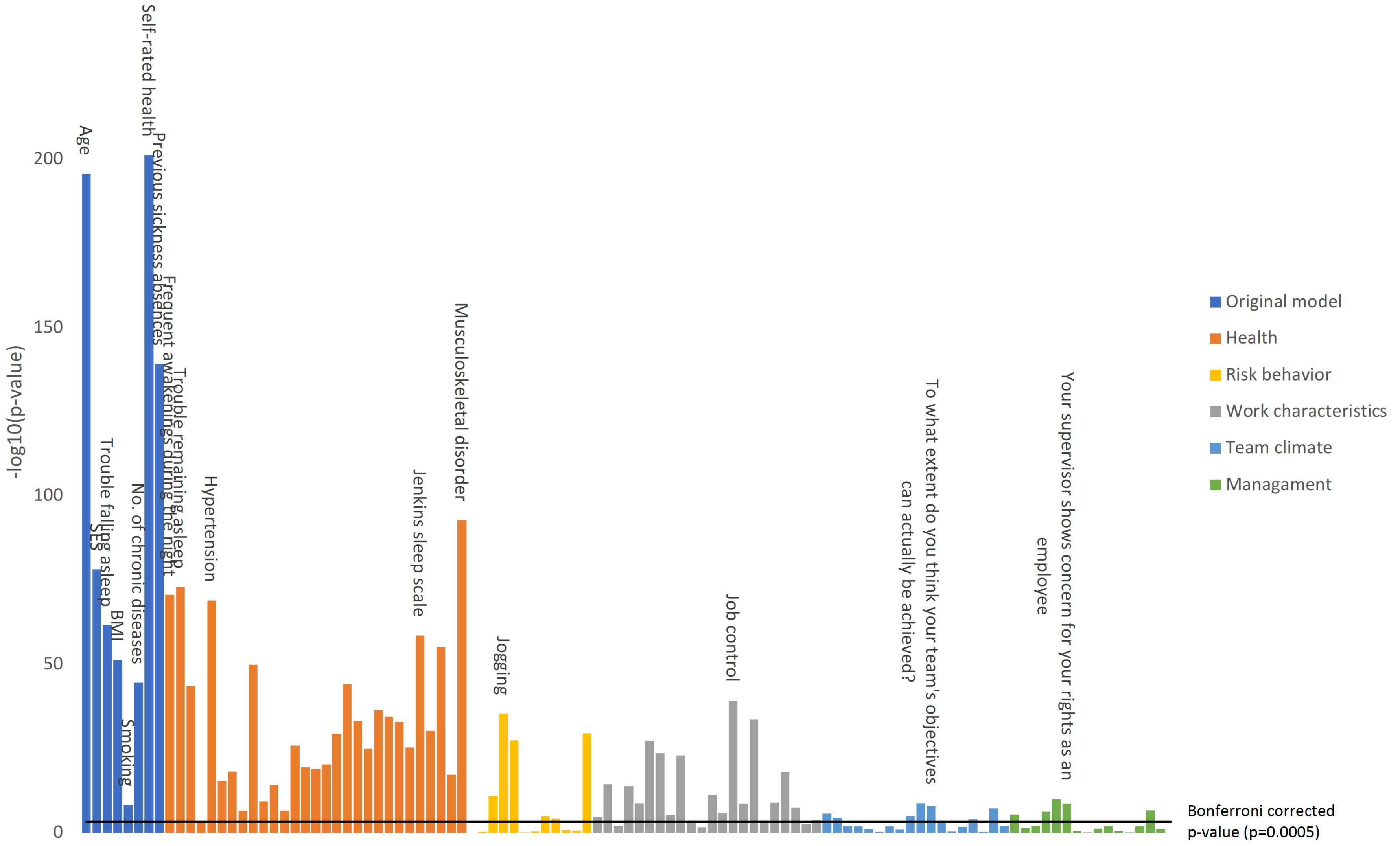


**Figure S4. Bivariate association between individual predictor items at baseline and work disability at follow-up in participants with respiratory disease at baseline**

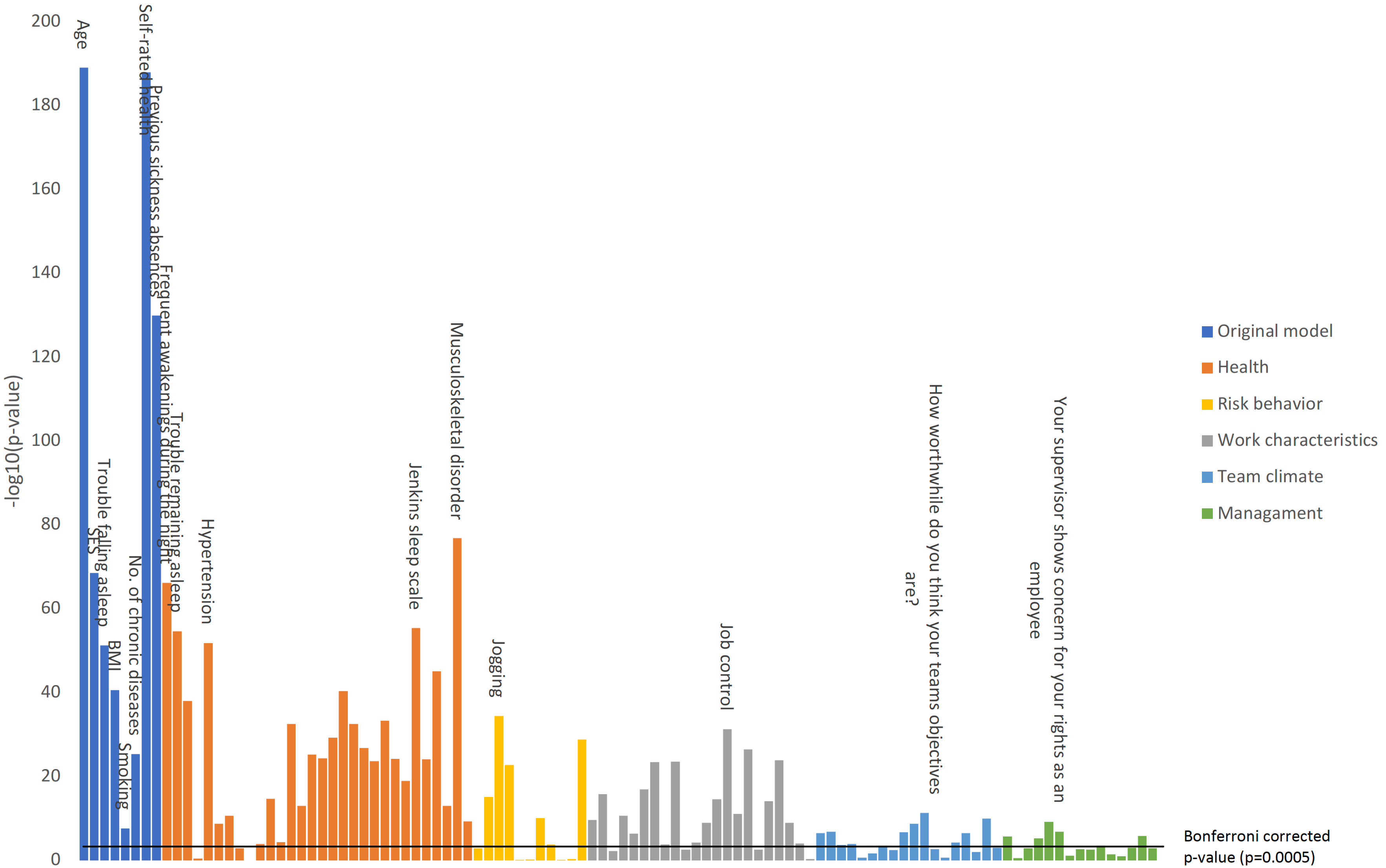


**Figure S5. Bivariate association between individual predictor items at baseline and work disability at follow-up in participants with depression at baseline**

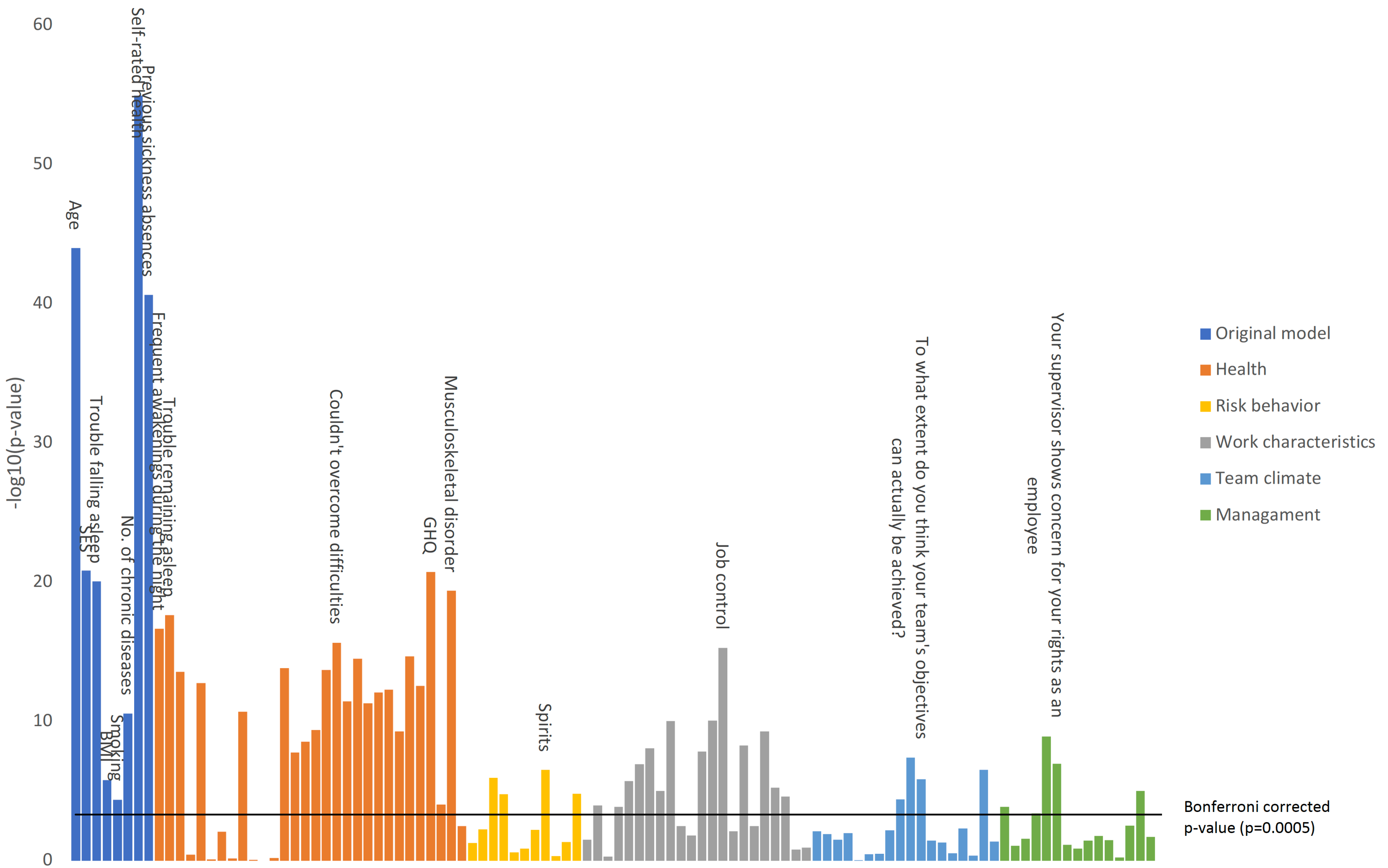


**Figure S6. Bivariate association between individual predictor items at baseline and work disability at follow-up in participants with diabetes at baseline**

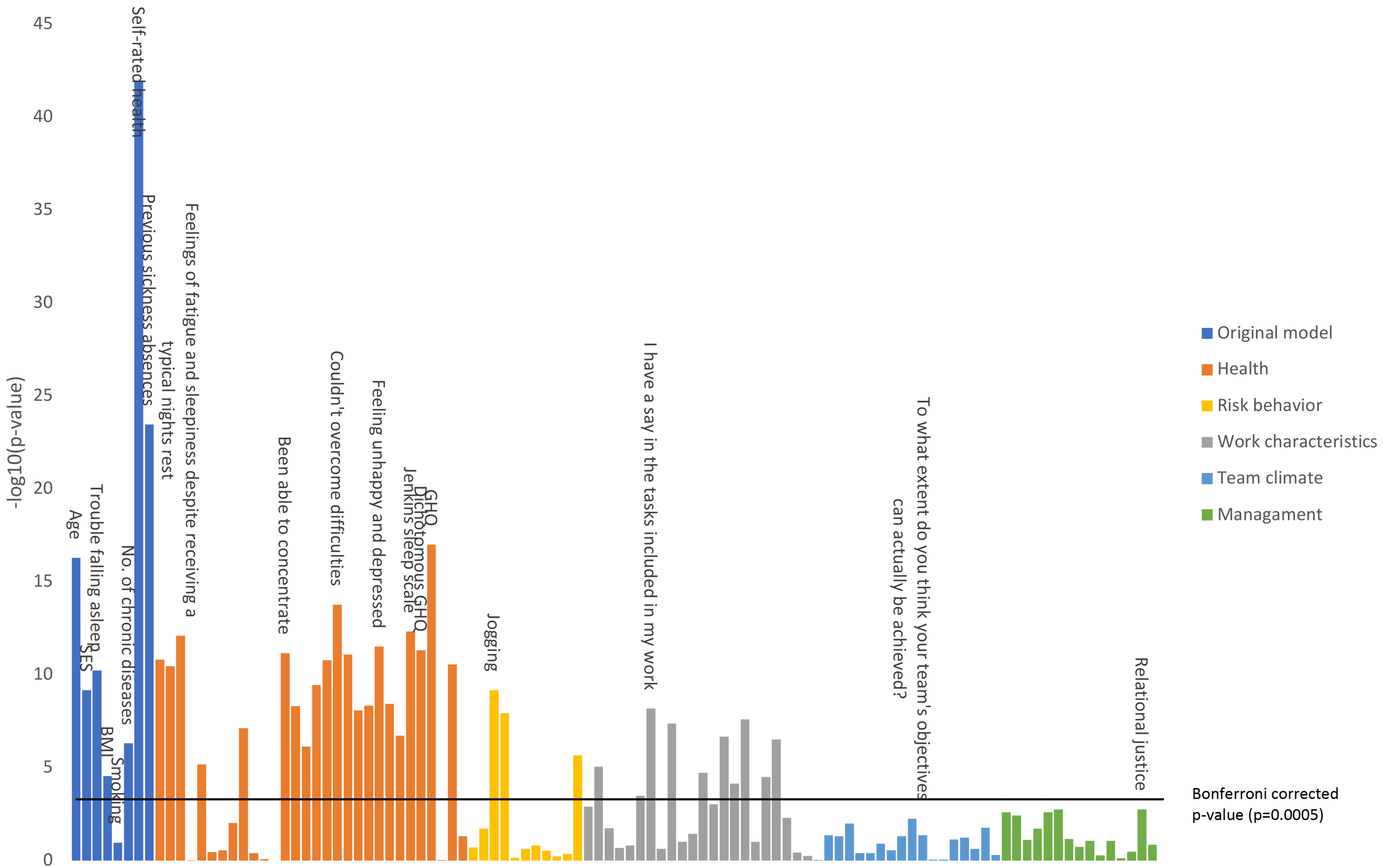


**Figure S7. Bivariate association between individual predictor items at baseline and work disability at follow-up in participants with cancer at baseline**

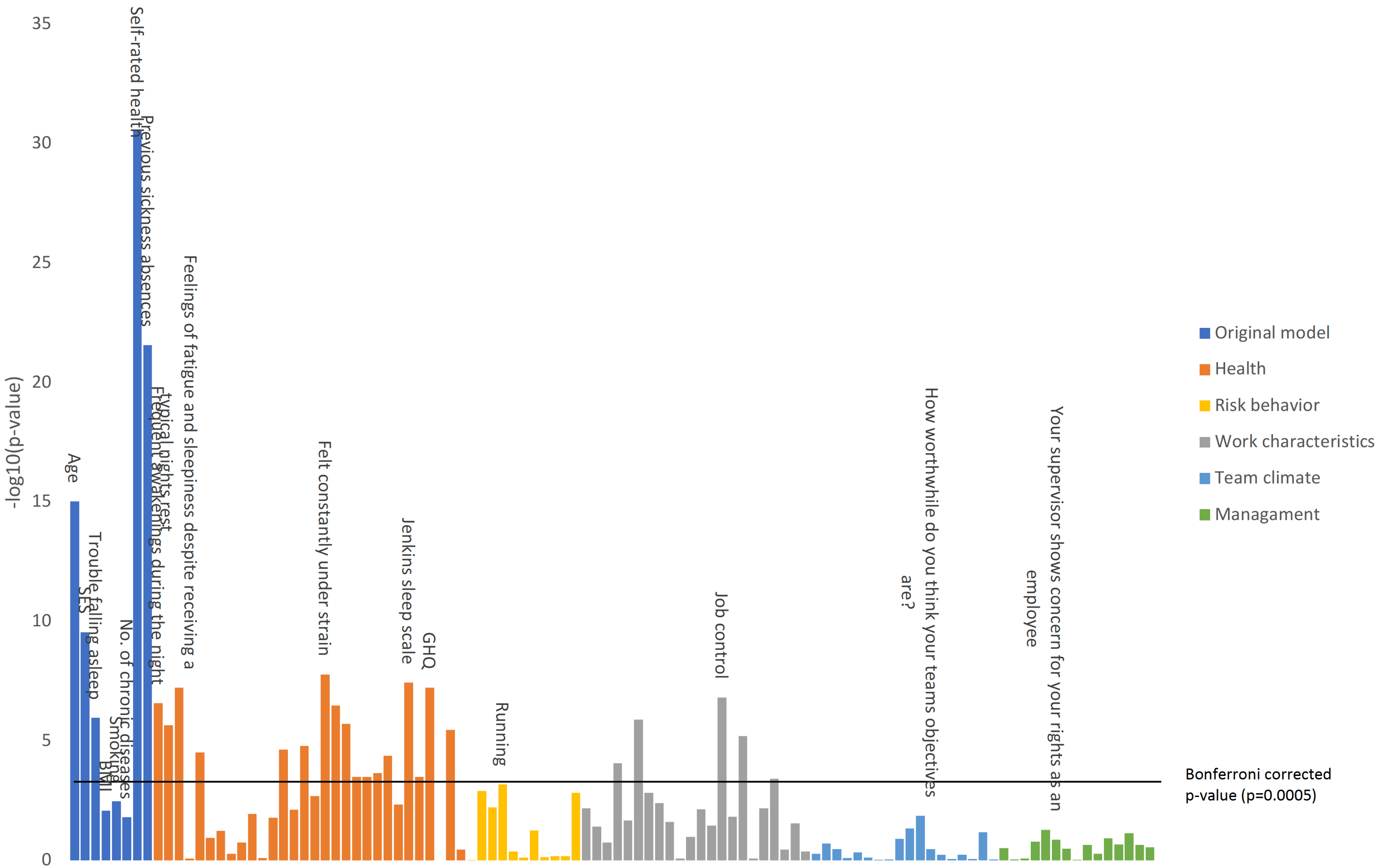


**Figure S8. Bivariate association between individual predictor items at baseline and work disability at follow-up in participants with coronary heart disease at baseline**

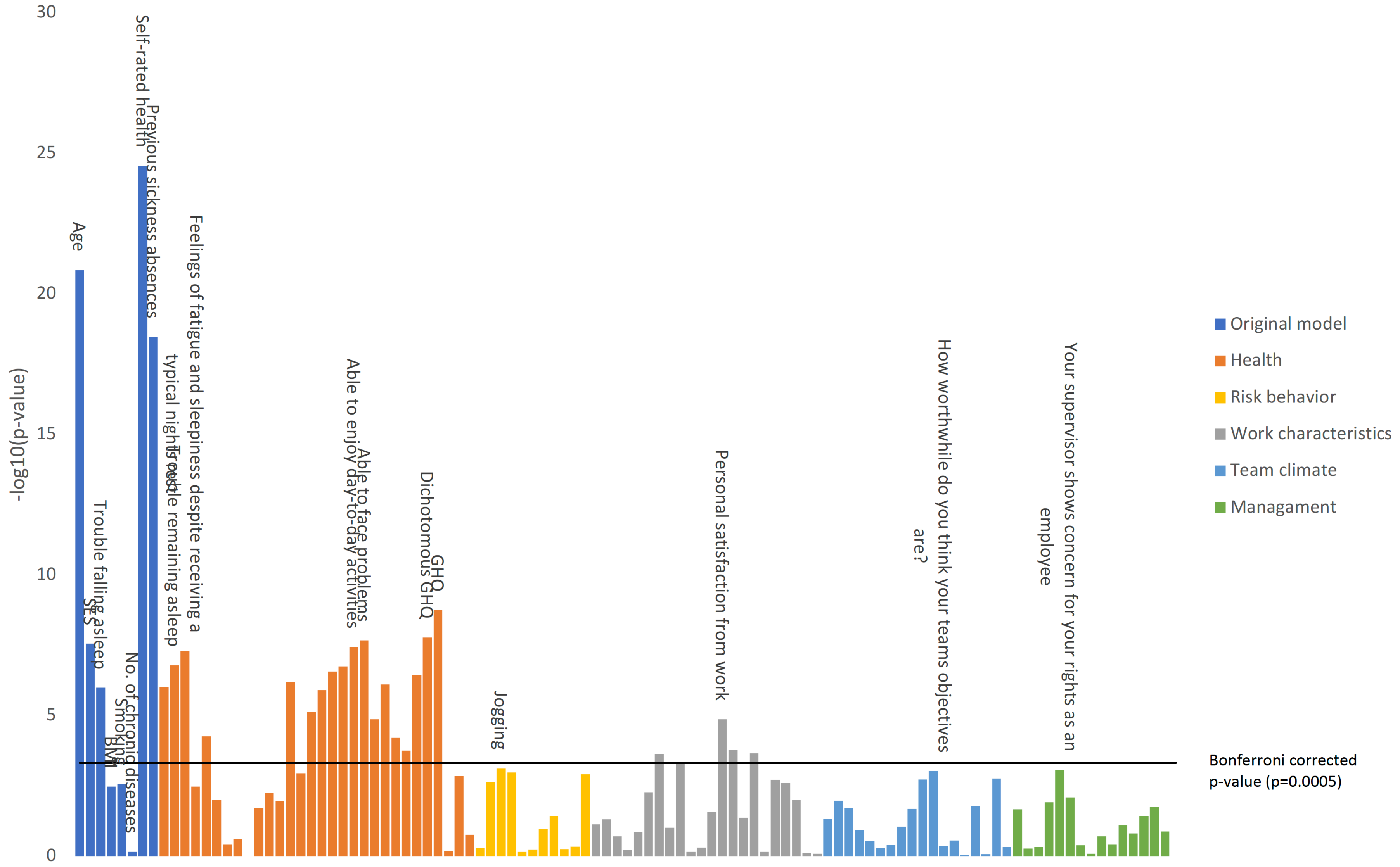


**Figure S9. Bivariate association between individual predictor items at baseline and work disability at follow-up in participants with comorbid depression and cardiometabolic disease (diabetes, CHD or stroke) at baseline**

### Table S4. Re-estimated coefficients and new predictive algorithms

| Disease | Variable | Classes | Coefficient/class 1 | 2 | 3 | 4 | 5 | 6 | 7 | Legend |
|---|---|---|---|---|---|---|---|---|---|---|
| MSD, C=0.80 (0.79, 0.81) | | | | | | | | | | |
| | Age | 1-6 | 0 | -0.2873 | -0.4692 | -1.0077 | -1.4558 | -1.671 | | 1='<35', 2='35-39', 3='40-44', 4='45-49', 5='50-54', 6='55+' |
| | BMI | 1-4 | -0.0542 | 0 | -0.076 | -0.1397 | | | | 1='<18.5', 2='18.5-<25', 3='25-<30', 4='30+' |
| | SES | 1-7 | 0 | -0.043 | -0.4308 | -0.3418 | -0.6404 | -0.6046 | -0.7126 | 1='Manager', 2=Senior specialist', 3='Specialist', 4='Office worker', 5='Service worker', 6='Process worker', 7='Other' |
| | Smoking | 1-2 | 0 | -0.124 | | | | | | 1='No', 2='Yes' |
| | Self-rated health | 1-5 | 0 | -0.2817 | -0.6463 | -1.1033 | -1.5051 | | | 1='Good', 2='Rather good', 3='Moderate', 4='Rather poor', 5='Poor' |
| | Number of sickness absences in previous year | 1-4 | 0 | -0.4247 | -0.6184 | -0.8252 | | | | 1='0', 2='1', 3='2', 4='3+' |
| | Self-reported depression | 1-2 | 0 | -0.2389 | | | | | | 1='No', 2='Yes' |
| | Effort* | 1-5 | 0 | 0.0075 | -0.1406 | -0.1483 | -0.2404 | | | Likert scale ranging from 1 = "very little" to 5 = "very much" |
| | Intercept / Log(Scale) | | 6.0473 / -0.3264 | | | | | | | |
| Migraine, C=0.83 (0.83, 0.84) | | | | | | | | | | |
| | Age | 1-6 | 0 | -0.2667 | -0.3559 | -0.8367 | -1.2783 | -1.4647 | | 1='<35', 2='35-39', 3='40-44', 4='45-49', 5='50-54', 6='55+' |
| | BMI | 1-4 | -0.0173 | 0 | -0.0696 | -0.153 | | | | 1='<18.5', 2='18.5-<25', 3='25-<30', 4='30+' |
| | SES | 1-7 | 0 | 0.0676 | -0.2944 | -0.2039 | -0.4635 | -0.3571 | -0.5776 | 1='Manager', 2=Senior specialist', 3='Specialist', 4='Office worker', 5='Service worker', 6='Process worker', 7='Other' |
| | Smoking | 1-2 | 0 | -0.1423 | | | | | | 1='No', 2='Yes' |
| | Number of chronic illness | 1-3 | 0 | -0.245 | -0.38 | | | | | 1='1', 2='2', 3='3+' |
| | Self-rated health | 1-5 | 0 | -0.1242 | -0.4803 | -0.9516 | -1.1781 | | | 1='Good', 2='Rather good', 3='Moderate', 4='Rather poor', 5='Poor' |
| | Number of sickness absences in previous year | 1-4 | 0 | -0.4321 | -0.6274 | -0.8612 | | | | 1='0', 2='1', 3='2', 4='3+' |
| | Self-reported MSD | 1-2 | 0 | -0.2403 | | | | | | 1='No', 2='Yes' |
| | Intercept / Log(Scale) | | 5.5881 / -0.3712 | | | | | | | |
| Hypertension, C=0.79 (0.79, 0.80) | | | | | | | | | | |
| | Age | 1-6 | 0 | -0.708 | -0.7202 | -1.1009 | -1.5534 | -1.7417 | | 1='<35', 2='35-39', 3='40-44', 4='45-49', 5='50-54', 6='55+' |
| | SES | 1-7 | 0 | -0.0674 | -0.3639 | -0.273 | -0.5827 | -0.558 | -0.7137 | 1='Manager', 2=Senior specialist', 3='Specialist', 4='Office worker', 5='Service worker', 6='Process worker', 7='Other' |
| | Number of chronic illness | 1-4 | 0 | -0.1912 | -0.3115 | -0.3957 | | | | 1='0', 2='1', 3='2', 4='3+' |
| | Self-rated health | 1-5 | 0 | -0.1254 | -0.4543 | -0.985 | -1.2665 | | | 1='Good', 2='Rather good', 3='Moderate', 4='Rather poor', 5='Poor' |
| | Number of sickness absences in previous year | 1-4 | 0 | -0.425 | -0.6817 | -0.734 | | | | 1='0', 2='1', 3='2', 4='3+' |
| | Shiftwork | 1-2 | 0 | -0.0799 | | | | | | 1='No', 2='Yes' |
| | Effort | 1-5 | 0 | -0.2776 | -0.2748 | -0.3743 | -0.4252 | | | Likert scale ranging from 1 = "very little" to 5 = "very much" |
| | Self-reported MSD | 1-2 | 0 | -0.2358 | | | | | | 1='No', 2='Yes' |
| | Intercept / Log(Scale) | | 6.3076 / -0.3527 | | | | | | | |
| Respiratory disease, C=0.82 (0.81, 0.83) | | | | | | | | | | |
| | Age | 1-6 | 0 | -0.3198 | -0.4333 | -0.8849 | -1.2447 | -1.4719 | | 1='<35', 2='35-39', 3='40-44', 4='45-49', 5='50-54', 6='55+' |
| | SES | 1-7 | 0 | -0.0195 | -0.456 | -0.4361 | -0.5588 | -0.5352 | -0.7523 | 1='Manager', 2=Senior specialist', 3='Specialist', 4='Office worker', 5='Service worker', 6='Process worker', 7='Other' |
| | Number of chronic illness | 1-4 | 0 | -0.1302 | -0.3148 | -0.4203 | | | | 1='0', 2='1', 3='2', 4='3+' |
| | Self-rated health | 1-5 | 0 | -0.3066 | -0.6458 | -1.121 | -1.4591 | | | 1='Good', 2='Rather good', 3='Moderate', 4='Rather poor', 5='Poor' |
| | Number of sickness absences in previous year | 1-4 | 0 | -0.4279 | -0.5175 | -0.7633 | | | | 1='0', 2='1', 3='2', 4='3+' |
| | Effort | 1-5 | 0 | -0.1467 | -0.0632 | -0.2482 | -0.3408 | | | Likert scale ranging from 1 = "very little" to 5 = "very much" |
| | Innovation / time for new ideas* | 1-5 | 0 | -0.0095 | 0.0142 | 0.1904 | 0.0914 | | | Likert scale ranging from 1 = "completely agree" to 5 = "completely disagree" |
| | Self-reported MSD | 1-2 | 0 | -0.2174 | | | | | | 1='No', 2='Yes' |
| | Intercept / Log(Scale) | | 6.0205 / -0.3435 | | | | | | | |
| Depression, C=0.78 (0.77, 0.78) | | | | | | | | | | |
| | Age | 1-6 | 0 | -0.0818 | -0.2438 | -0.7123 | -1.0733 | -1.3342 | | 1='<35', 2='35-39', 3='40-44', 4='45-49', 5='50-54', 6='55+' |
| | BMI | 1-4 | -0.0536 | 0 | -0.0831 | -0.196 | | | | 1='<18.5', 2='18.5-<25', 3='25-<30', 4='30+' |
| | SES | 1-7 | 0 | 0.0458 | -0.2559 | -0.2574 | -0.4078 | -0.3966 | -0.5348 | 1='Manager', 2=Senior specialist', 3='Specialist', 4='Office worker', 5='Service worker', 6='Process worker', 7='Other' |
| | Number of chronic illness | 1-3 | 0 | -0.0697 | -0.2303 | | | | | 1='1', 2='2', 3='3+' |
| | Self-rated health | 1-5 | 0 | -0.1492 | -0.471 | -0.9069 | -1.2568 | | | 1='Good', 2='Rather good', 3='Moderate', 4='Rather poor', 5='Poor' |
| | Difficulty falling asleep | 1-6 | 0 | -0.043 | -0.0966 | -0.1399 | -0.0956 | -0.2353 | | 1='Never', 2='1-3 nights/month', 3='once/week', 4='2-4 nights per week"', 5='5-6 nights per week', 6='Almost every night' |
| | Number of sickness absences in previous year | 1-4 | 0 | -0.3488 | -0.5904 | -0.9371 | | | | 1='0', 2='1', 3='2', 4='3+' |
| | Self-reported MSD | 1-2 | 0 | -0.151 | | | | | | 1='No', 2='Yes' |
| | Intercept / Log(Scale) | | 5.3043 / -0.2333 | | | | | | | |

| | | | | | | | | | | | |
|---|---|---|---|---|---|---|---|---|---|---|---|
| **Diabetes, C=0.79 (0.77, 0.81)** | | | | | | | | | | | |
| | Age | 1-6 | | 0 | -0.4991 | -0.622 | -0.9117 | -1.4321 | -1.4612 | | 1='<35', 2='35-39', 3='40-44', 4='45-49', 5='50-54', 6='55+' |
| | SES | 1-7 | | 0 | 0.0428 | -0.245 | -0.1714 | -0.3837 | -0.4702 | -0.5859 | 1='Manager', 2=Senior specialist', 3='Specialist', 4='Office worker', 5='Service worker', 6='Process worker', 7='Other' |
| | Self-rated health | 1-5 | | 0 | -0.4466 | -0.8065 | -1.3214 | -1.4737 | | | 1='Good', 2='Rather good', 3='Moderate', 4='Rather poor', 5='Poor' |
| | Number of sickness absences in previous year | 1-4 | | 0 | -0.4449 | -0.7388 | -0.6014 | | | | 1='0', 2='1', 3='2', 4='3+' |
| | Reward / personal satisfaction | 1-5 | | 0 | 0.1448 | 0.0096 | -0.1635 | -0.1707 | | | Likert scale ranging from 1 = "very much" to 5 = "very little" |
| | Innovation / co-operation for new ideas* | 1-5 | | 0 | -0.2719 | -0.1255 | 0.0177 | 0.0754 | | | Likert scale ranging from 1 = "completely agree" to 5 = "completely disagree" |
| | Self-reported depression | 1-2 | | 0 | -0.2912 | | | | | | 1='No', 2='Yes' |
| | Self-reported MSD | 1-2 | | 0 | -0.1576 | | | | | | 1='No', 2='Yes' |
| | Intercept / Log(Scale) | | 5.8554 / -0.2743 | | | | | | | | |
| **Cancer, C=0.74 (0.72, 0.76)** | | | | | | | | | | | |
| | Age | 1-6 | | 0 | 0.7137 | -0.2989 | -0.2182 | -0.5742 | -0.7943 | | 1='<35', 2='35-39', 3='40-44', 4='45-49', 5='50-54', 6='55+' |
| | SES | 1-7 | | 0 | 0.0786 | 0.0184 | 0.0565 | -0.2462 | -0.0411 | -0.2568 | 1='Manager', 2=Senior specialist', 3='Specialist', 4='Office worker', 5='Service worker', 6='Process worker', 7='Other' |
| | Self-rated health | 1-5 | | 0 | -0.464 | -0.7688 | -1.388 | -2.1199 | | | 1='Good', 2='Rather good', 3='Moderate', 4='Rather poor', 5='Poor' |
| | Number of sickness absences in previous year | 1-4 | | 0 | -0.4614 | -0.5213 | -0.9575 | | | | 1='0', 2='1', 3='2', 4='3+' |
| | Task orientation / weaknesses | | | 0 | 0.2875 | 0.3744 | 0.4419 | 0.4931 | | | Likert scale ranging from 1 = "very much" to 5 = "very little" |
| | Job control / a lot of say | 1-5 | | 0 | -0.0219 | -0.207 | -0.2906 | -0.3205 | | | Likert scale ranging from 1 = "completely agree" to 5 = "completely disagree" |
| | Job control / little freedom | 1-5 | | 0 | -0.0248 | 0.122 | 0.1797 | 0.3082 | | | Likert scale ranging from 1 = "completely agree" to 5 = "completely disagree" |
| | Self-reported MSD | 1-2 | | 0 | -0.2083 | | | | | | 1='No', 2='Yes' |
| | Intercept / Log(Scale) | | 4.7732 / -0.1273 | | | | | | | | |
| **CHD, C=0.77 (0.74, 0.79)** | | | | | | | | | | | |
| | Age | 1-6 | | 0 | 0.8087 | 0.7104 | -0.4283 | -0.9428 | -0.9543 | | 1='<35', 2='35-39', 3='40-44', 4='45-49', 5='50-54', 6='55+' |
| | SES | 1-7 | | 0 | 0.318 | 0.0684 | -0.011 | -0.0548 | -0.1018 | -0.367 | 1='Manager', 2=Senior specialist', 3='Specialist', 4='Office worker', 5='Service worker', 6='Process worker', 7='Other' |
| | Number of chronic illness | 1-3 | | 0 | -0.1758 | -0.2125 | | | | | 1='1', 2='2', 3='3+' |
| | Self-rated health | 1-5 | | 0 | -0.8038 | -1.0533 | -1.5925 | -1.8411 | | | 1='Good', 2='Rather good', 3='Moderate', 4='Rather poor', 5='Poor' |
| | Number of sickness absences in previous year | 1-4 | | 0 | -0.3633 | -0.7461 | -0.5799 | | | | 1='0', 2='1', 3='2', 4='3+' |
| | Self-reported hypertension | 1-2 | | 0 | -0.202 | | | | | | 1='No', 2='Yes' |
| | Self-reported MSD | 1-2 | | 0 | -0.2329 | | | | | | 1='No', 2='Yes' |
| | Self-reported respiratory disease | 1-2 | | 0 | 0.1749 | | | | | | 1='No', 2='Yes' |
| | Intercept / Log(Scale) | | 5.4092 / -0.2257 | | | | | | | | |
| **DCD, C=0.78 (0.75, 0.80)** | | | | | | | | | | | |
| | Age | 1-6 | | 0 | -0.0598 | -0.1713 | -0.4495 | -1.0428 | -1.1964 | | 1='<35', 2='35-39', 3='40-44', 4='45-49', 5='50-54', 6='55+' |
| | SES | 1-7 | | 0 | 0.4618 | 0.2905 | 0.4255 | 0.1116 | 0.2913 | -0.0872 | 1='Manager', 2=Senior specialist', 3='Specialist', 4='Office worker', 5='Service worker', 6='Process worker', 7='Other' |
| | Smoking | 1-2 | | 0 | -0.2495 | | | | | | 1='No', 2='Yes' |
| | Self-rated health | 1-5 | | 0 | 0.0365 | -0.566 | -1.1546 | -1.3794 | | | 1='Good', 2='Rather good', 3='Moderate', 4='Rather poor', 5='Poor' |
| | Number of sickness absences in previous year | 1-4 | | 0 | -0.6047 | -0.7199 | -0.9259 | | | | 1='0', 2='1', 3='2', 4='3+' |
| | Self-reported hypertension | 1-2 | | 0 | -0.2578 | | | | | | 1='No', 2='Yes' |
| | Innovation /problems* | 1-5 | | 0 | 0.1225 | 0.0802 | 0.3537 | 0.4378 | | | Likert scale ranging from 1 = "completely agree" to 5 = "completely disagree" |
| | Self-reported allergic rhinitis | 1-2 | | 0 | 0.1979 | | | | | | 1='No', 2='Yes' |
| | Intercept / Log(Scale) | | 4.4099 / -0.1529 | | | | | | | | |

Effort: "How much do you feel you invest in your job in terms of skill and energy?"
Innovation / time for new ideas: "In this team we take the time needed to develop new ideas"
Reward / personal satisfaction: "Do you feel that you get personal satisfaction of your work?"
Innovation / co-operation for new ideas: 'People in the team co-operate in order to help develop and apply new ideas'
Innovation / problems: 'People in this team are always searching for fresh, new ways of looking at problems'
Job control / a lot of say: 'I have a lot of say in the tasks included in my work'
Job control / little freedom: 'I have very little freedom to decide how I do my work'
Task orientation / weaknesses: 'Does the team critically appraise potential weaknesses in what it is doing in order to achieve the best possible outcome?'

### Table S5. Sensitivity, specificity and positive and negative predictive value for the existing prediction model in all participants and subgroups of individuals with no history of sickness absence and those with a chronic condition at baseline

| Predictive performance for a positive test | Cut-off (%) for a positive test result | | | | | | |
|---|---|---|---|---|---|---|---|
| | 5 | 10 | 20 | 30 | 40 | 50 | 60 |
| **Musculoskeletal disorders** | | | | | | | |
| Sensitivity | 95.3 | 86.6 | 65.9 | 46.6 | 32.7 | 21.1 | 12.8 |
| Specificity | 29.8 | 49.6 | 73.4 | 86.4 | 92.9 | 96.4 | 98.3 |
| Positive predictive value | 18.2 | 22.0 | 28.8 | 36.0 | 43.2 | 48.9 | 54.6 |
| Negative predictive value | 97.5 | 95.8 | 92.9 | 90.8 | 89.4 | 88.2 | 87.3 |
| **Migraine** | | | | | | | |
| Sensitivity | 92.0 | 83.1 | 63.6 | 47.6 | 33.3 | 22.1 | 14.3 |
| Specificity | 45.8 | 63.0 | 80.7 | 90.0 | 94.8 | 97.4 | 98.8 |
| Positive predictive value | 15.7 | 19.8 | 26.6 | 34.5 | 41.4 | 48.5 | 57.3 |
| Negative predictive value | 98.1 | 97.1 | 95.3 | 94.0 | 92.8 | 91.9 | 91.3 |
| **Hypertension** | | | | | | | |
| Sensitivity | 96.4 | 89.6 | 71.2 | 53.2 | 37.3 | 24.8 | 15.2 |
| Specificity | 19.5 | 38.3 | 65.6 | 82.1 | 90.6 | 95.1 | 97.8 |
| Positive predictive value | 17.6 | 20.6 | 26.9 | 34.6 | 41.5 | 47.5 | 55.0 |
| Negative predictive value | 96.8 | 95.4 | 92.8 | 90.8 | 89.0 | 87.7 | 86.6 |
| **Respiratory disease** | | | | | | | |
| Sensitivity | 93.0 | 84.8 | 66.0 | 49.9 | 35.8 | 24.1 | 15.1 |
| Specificity | 40.1 | 57.1 | 76.8 | 87.1 | 92.8 | 96.2 | 98.2 |
| Positive predictive value | 17.8 | 21.6 | 28.4 | 35.0 | 41.2 | 47.1 | 54.2 |
| Negative predictive value | 97.6 | 96.4 | 94.2 | 92.6 | 91.2 | 90.1 | 89.2 |
| **Depression** | | | | | | | |
| Sensitivity | 93.4 | 86.2 | 69.7 | 53.7 | 40.1 | 27.4 | 18.0 |
| Specificity | 27.7 | 45.7 | 68.3 | 81.5 | 89.3 | 93.9 | 97.0 |
| Positive predictive value | 20.5 | 24.1 | 30.6 | 36.7 | 42.9 | 47.3 | 54.3 |
| Negative predictive value | 95.4 | 94.3 | 91.8 | 89.8 | 88.2 | 86.6 | 85.5 |
| **Diabetes** | | | | | | | |
| Sensitivity | 97.0 | 91.6 | 77.7 | 62.4 | 46.3 | 32.8 | 20.7 |
| Specificity | 19.2 | 32.0 | 53.7 | 72.4 | 83.9 | 91.4 | 95.7 |
| Positive predictive value | 19.1 | 20.9 | 24.8 | 30.7 | 36.1 | 42.8 | 48.7 |
| Negative predictive value | 97.0 | 95.1 | 92.5 | 90.8 | 88.8 | 87.4 | 86.0 |
| **Cancer** | | | | | | | |
| Sensitivity | 93.1 | 80.4 | 58.3 | 42.2 | 30.3 | 18.9 | 13.2 |
| Specificity | 21.2 | 40.7 | 66.5 | 81.2 | 90.3 | 95.7 | 97.9 |
| Positive predictive value | 16.4 | 18.3 | 22.4 | 27.1 | 34.1 | 42.3 | 51.1 |
| Negative predictive value | 94.9 | 92.6 | 90.6 | 89.5 | 88.7 | 87.7 | 87.2 |
| **Coronary heart disease** | | | | | | | |
| Sensitivity | 99.2 | 97.2 | 88.9 | 76.1 | 62.8 | 47.2 | 31.7 |
| Specificity | 5.6 | 12.5 | 33.6 | 54.5 | 69.7 | 81.0 | 89.7 |
| Positive predictive value | 23.5 | 24.5 | 28.1 | 32.8 | 37.7 | 42.1 | 47.4 |
| Negative predictive value | 96.3 | 94.0 | 91.2 | 88.7 | 86.5 | 84.0 | 81.8 |
| **Comorbid depression and cardiometabolic disease** | | | | | | | |
| Sensitivity | 98.1 | 96.3 | 90.7 | 79.8 | 67.7 | 52.8 | 38.5 |
| Specificity | 11.0 | 20.1 | 38.1 | 54.8 | 67.7 | 79.9 | 88.2 |
| Positive predictive value | 29.7 | 31.6 | 36.0 | 40.3 | 44.6 | 50.1 | 55.6 |
| Negative predictive value | 93.9 | 93.4 | 91.4 | 87.6 | 84.5 | 81.5 | 78.9 |
| **Low-risk population** | | | | | | | |
| Sensitivity | 81.6 | 63.5 | 35.0 | 17.2 | 8.3 | 3.5 | 1.2 |
| Specificity | 61.8 | 78.5 | 92.3 | 97.3 | 99.1 | 99.6 | 99.9 |
| Positive predictive value | 11.0 | 14.5 | 20.8 | 26.8 | 33.8 | 34.9 | 38.6 |
| Negative predictive value | 98.3 | 97.4 | 96.1 | 95.3 | 94.9 | 94.7 | 94.6 |
| **Total cohort** | | | | | | | |
| Sensitivity | 87.9 | 75.0 | 52.5 | 35.9 | 24.0 | 15.0 | 9.1 |
| Specificity | 56.3 | 73.0 | 88.0 | 94.3 | 97.2 | 98.6 | 99.4 |
| Positive predictive value | 14.4 | 18.9 | 26.7 | 34.6 | 41.9 | 48.0 | 55.3 |
| Negative predictive value | 98.2 | 97.2 | 95.7 | 94.6 | 93.9 | 93.3 | 92.9 |

**Statistical code**

**Imputation of missing data**

```
imputed <- mice(idata, m=1, method = "pmm", seed=1)
idata_all <-complete(imputed,1)
```

**C-index**

```
#Create survival object
sv <- Surv(data$tkseuraika, data$tkstatus)

#Calculate c-index
c_old <- rcorr.cens(data$predicts_old, sv)
old_lci <- round(c_old["C Index"]-1.96*(c_old["S.D."]/2),4)
old_hci <- round(c_old["C Index"]+1.96*(c_old["S.D."]/2),4)
```

**Re-estimating the coefficients of the original model**

```
#Define the model
formodel <- as.formula("Surv(tkseuraika, tkstatus) ~ as.factor(agecat)+
                              as.factor(bmi00)+
                              as.factor(ses)+
                              as.factor(smoking)+
                              as.factor(illnesses)+
                              as.factor(srh00)+
                              as.factor(sleep0)+
                              as.factor(sickabs)")
#Estimate new model
newmodel <- psm(formodel, data=data, dist="lognormal", x=T, y=T)
newmodel
```

**An example of selecting a completely new set of predictors**

```
#Create a formula vector to be used in redundancy analysis
forred <- as.formula(paste("~",paste0(allvars,collapse="+")))
forred
#Redundancy analysis
red <- redun(forred, nk=0, data=idata)
red

#Delete redundant variables from "allvars" and create a new vector called "vars_subset"
#that is used from here on out.
vars_subset <- allvars

for(d in 1:length(red$Out)){
  vars_subset <- vars_subset[vars_subset !=red$Out[d]]
```

```
}
vars_subset


######### Predictor selection (stepwise backward elimination) ###########

#First we fit a full parametric survival model
  #Define the model
  forwei <- as.formula(paste("Surv(", "tkseuraika",",","tkstatus",") ~", paste(vars_subset,
collapse='+')))
  #Run the analysis
  fullWei <- psm(forwei, data=idata, dist = "weibull")
fullWei
#Add linear predictor values from full Weibull model to dataframe
idata$weilp <- predict(fullWei, type="lp")

#Then an OLS model with the outcome from Weibull model as dependent variable
  #Define the model
  forols <- as.formula(paste("weilp ~", paste0(vars_subset, collapse='+')))
  #Run the predictor selection
  model = regsubsets(forols, data = idata,  method = "backward", nvmax=50)
  #Save summary of the results to "modelsum" (used to extract predictor names)
  modelsum <-summary(model)

nvars <- 8

#Extract variable names from "modelsum"
i<-2
iv<-vector()
modelnames <- colnames(modelsum$which[])
while(i<length(modelsum$which[nvars,])+1){
  if(modelsum$which[nvars,i]==1)
     iv <- c(iv,paste(modelnames[i]))
    i<-i+1
     }

##Define a formula from extracted variables
finalmodel <- as.formula(paste("Surv(tkseuraika, tkstatus) ~",paste(iv, collapse =  "+")))
finalmodel

##Add predicted values to dataframe from the model derived from backward elimination
weimodel<-psm(finalmodel, data=idata, dist="weibull", x=T, y=T)
weimodel
idata$predicts <- predict(weimodel, type="lp")


idata$agecat <- as.numeric(cut(idata$age,c(15,35,40,45,50,55,70), right = F))
```

```
idata$bmicat  <- as.numeric(cut(idata$bmi00,c(15,18.5,25,30,55), right = F))

idata$ sickabs_cat <- ifelse(idata$ sickabs>3,3,idata$ sickabs)

finalmodel <- as.formula(paste("Surv(tkseuraika, tkstatus) ~ as.factor(agecat) +
as.factor(K31_2) +
                         as.factor(ses) + as.factor(sickabs_cat) + as.factor(eff00) +
as.factor(tautisum00) +
                        as.factor(ttila00) + as.factor(k8y8_9_11) "))


##Add predicted values to dataframe from the model derived from backward elimination
weimodel<-psm(finalmodel, data=idata, dist="weibull", x=T, y=T)
weimodel
idata$predicts <- predict(weimodel, type="lp")
```
